# Warning system for Extreme weather events, Awareness Technology for Healthcare, Equitable delivery, and Resilience (WEATHER) Project: A mixed methods research study protocol

**DOI:** 10.1101/2025.01.14.25320537

**Authors:** Mary Lynch, Fiona Harris, Michelle Ierna, Ozayr Mahomed, Fiona Henriquez-Mui, Michael Gebreslasie, David Ndzi, Serestina Viriri, Muhammad Zeeshan Shakir, Natalie Dickinson, Caroline Miller, Nisha Nadesan-Reddy, Fikile Nkwanyana, Llinos Haf Spencer, Saloshni Naidoo

**Author notes:** **Corresponding author 1:** Professor Mary Lynch, Vice Dean for Research, Faculty of Nursing and Midwifery, Royal College of Surgeons, Ireland, 123 St Stephen’s Green, Dublin, County Dublin, D02 YN77, Ireland. **Corresponding author 2:** Dr Llinos Haf Spencer, Research Fellow, Faculty of Nursing and Midwifery, Royal College of Surgeons, Ireland, 123 St Stephen’s Green, Dublin, County Dublin, D02 YN77, Ireland.

## Abstract

**Background:** Extreme weather events (EWEs) in KwaZulu-Natal province, South Africa are increasingly common causing disease outbreaks, loss of homes, damage to infrastructure, increased mortality and life as well as increasing the pressure on the health system. An Early Warning System (EWS) intervention is needed to alert communities and government increasing preparedness for floods, improving resilience of health services to predict, detect and respond to emergencies and disease outbreaks. The aim of this mixed methods study is to develop, implement and evaluate an EWS as an alert system for communities and government agencies to detect, identify and prevent disease outbreaks strengthening the health system response.

**Methods:** The study sites are eThekwini and Ugu, municipalities in KwaZulu-Natal province, South Africa and will comprise the selected community and health facilities in selected sites in these two health districts. This transdisciplinary project will build on prior engagement with communities/health providers (e.g., communities, professionals, policy makers) integrating a participatory, co-creation approach. To build the foundation for this study and understand the impact of climate change on disease outbreaks, a systematic review will be conducted. We will also design, develop and evaluate the performance of an EWS and will link data on the two (rural and urban) healthcare and environmental contexts and integrate EWS for change, clarify how these will be delivered for vulnerable communities, and test and adapt the EWS. Following, an assessment of the disease burden at primary health care clinics in Ugu and eThekwini will be undertaken.

Subsequently, an examination of the health needs along with community experiences, during, an EWE, as well as assessment of the health systems resilience in responding to an EWE. Finally, an evaluation will be conducted on the design, development and evaluation of the performance of an EWS intervention and an evaluation and costing of the implementation of the EWS. Ethics for the project will be sought from the relevant ethical committees and informed consent taken by voluntary participants in the study.

**Discussion:** Based on the study findings, recommendations will be made to the Department of Health with respect to an early warning system and health systems resilience.

**Trial registration:** Not applicable

**Funding:** This study is funded by funded by a Research and Innovation for Global Health Transformation National Institute for Health and Care Research (NIHR) award (NIHR204825).

## Background

South Africa (SA) is susceptible to extreme weather events (EWEs) linked to climate change. These EWEs have included floods, droughts, fires and large storms [1] which caused socioeconomic disruption and damage with an estimated cost of USD 10 billion between 1980 and 2015 [2].

KwaZulu-Natal (KZN) province has a subtropical climate with hot humid summers and dry winters and an annual rainfall of 1,009 millimetres. As an example, the rainfall on Monday 11th April 2022 was equivalent ∼30% of the annual mean falling over a period of two days. Flooding associated with this rainfall event resulted in the displacement of over 40,000 people, more than 500 deaths, and reports of dozens of missing people [3]. The catastrophic damage to regional infrastructure led to power, water and food not being available to many residents during and after the flooding. Many areas in the province also suffered extensive property losses, including the washing away of roads, destruction of bridges, and massive damage to around 12,000 homes causing severe flood damage leading to limited access to safe, clean drinking water, and outbreaks of illnesses such as diarrhoea [3]. In June 2024 some of the flood effected towns (e.g. Tongaat) in the North of Durban were struck by a tornado, which caused further severe damage, including damage to roofs and walls, uprooting of trees and large vehicles being blown from their original position [4]. Climate change is affecting health in several ways, resulting in increased morbidity and mortality in countries around the world. This is owing to an increase in EWEs which are affecting food systems, increasing contamination of water sources and increasing water and food-borne illnesses [2]. This has disproportionately affected the vulnerable: pregnant women, children and the elderly in already poverty-stricken communities. Several of the social determinants of health have been further undermined, including access to health care. The increased prevalence of climate linked disease in South Africa is having a major impact on an already resource constrained health system [5].

Diarrhoea is one of the leading causes of morbidity and mortality in children under-five in South Africa, with official statistics from Statistics South Africa estimating that diarrhoea accounts for approximately 20% of under-five deaths [6]. Poor nutritional status, poor environmental conditions, and illnesses such as Human Immunodeficiency Virus (HIV)/ Acquired Immune Deficiency Syndrome (AIDS). HIV/AIDS make children more susceptible to 6 severe diarrhoea and dehydration [7].

Episodes of persistent diarrhoea also worsen a child’s condition and nutritional status due to decreased food intake and nutrient absorption [7]. In HIV-infected children, persistent diarrhoea is associated with an 11-fold increase in mortality [7]. Diarrhoea is closely linked to socio-economic status and has the most adverse effects in South Africa’s impoverished communities [8].

In addition, to the impact on the individual and communities, EWEs cause health service disruptions in terms of access. It is therefore necessary to build resilient health systems to ensure effective disaster risk management and the delivery of quality healthcare [9]. The World Health Organization (WHO) defines a climate resilient health system as one that can anticipate, respond to, cope with, recover from and adapt to climate-related adverse events [10], [11]. Through this response, a sustained improvement in overall planetary health is ensured despite the adverse climatic conditions. To ensure this resilient health system, the building blocks (i.e. leadership and governance, health workforce, health information systems, essential medical products and technologies, service delivery, and financing of the health system) must be made climate resilient [12]. A climate resilient health system, in turn, must support the strengthening of community engagement and community empowerment to build local capacity and resilience.

To mitigate the impact of flooding due to EWEs impact on human health and healthcare delivery, a **W**arning system for **E**WEs, that is established with particular attention given to **A**wareness **T**echnology for **H**ealthcare, **E**quitable delivery, and **R**esilience (WEATHER) is essential. The WEATHER project will inform upstream resource management for effective/efficient action to alleviate EWEs related health risks whilst facilitating health system strengthening and response.

### Overall aim

The overall aim of the WEATHER project is to develop, implement and evaluate an early warning system (EWS) intervention as a local alert system for communities and government to detect, identify and prevent disease outbreaks strengthening the health systems response.

### Overall objectives

1. Develop a community engagement and co-creation strategy informing Objectives 2-7.
2. Assess the relationship between EWEs, health risk agents and reported disease outbreaks.
3. Map/review healthcare delivery organisations to establish capacity and training needs.
4. Develop an EWS and pathogen and contamination management tool.
5. Support and deliver disaster management and intervention implementation training to health professionals.
6. Evaluate the impact of the EWS on EWE preparedness, system resilience and establish cost- effectiveness.
7. Build local research capability, strengthen knowledge transfer with other lower- and middle-income countries.

## Methods

This transdisciplinary project will build on prior engagement with communities/health providers (e.g communities, professionals, policy makers) integrating a participatory, co-creation approach. An over-arching intervention development framework will guide this project with evaluation conducted via a longitudinal, mixed methods approach in 4 Work Packages (WPs). These work packages will include a systematic review, assessment of the burden of disease in four selected communities in South Africa. WP2 will deploy and assess a low-cost advanced EWS in EWEs. WP3 will develop and deliver training in disaster management to health professionals for improving health systems resilience. WP4 will apply an overarching Realist Evaluation and Social Cost Benefit Analysis (SCBA) to evaluate the efficiency, acceptability and cost effectiveness of EWS impact on health outcomes during flooding in KZN province. These work packages will be described in more detail below:

### Work package 1: Systematic review

#### Aim

This WP will examine the evidence on EWEs and infectious disease outbreaks which will inform a comprehensive framework for monitoring health conditions and types of infections to build evaluation endpoint for the EWS. Design and development of a pathogen and contamination management tool in WP1 which will share information with EWS development in WP2.

#### Objectives

A comprehensive search strategy will be developed using appropriate key words, Medical Subject Headings (MeSH) and free text terms to maximize the retrieval of potentially relevant studies. The search will be conducted across various electronic databases without a date limit: Medline/PubMed, EBSCO (PsycINFO and CINAHL), Cochrane libraries, EMBASE, Web of Science and WHO Regional Databases. Screening and eligibility determination using a two-reviewer system (with consensus for disagreements and conferral with a third-party adjudicator if a consensus was unable to be reached), articles will be identified and screened by reviewing the title and abstract to remove all articles that clearly did not meet the eligibility criteria. Details about the included and excluded studies will be presented in a PRISMA diagram [13], [14]. The full text of the remaining articles will be reviewed by two reviewers, any ensuing discrepancies will be resolved by discussion or the involvement of the third reviewer if a consensus is not reached by the first two reviewers.

Eligibility will be based on the following inclusion and exclusion criteria:

##### Inclusion criteria

Literature with substantial focus on flooding and intervention models including peer-reviewed journal articles, systematic reviews, scoping reviews, meta-analysis and rapid reviews, government and non-governmental organisation reports and academic dissertations, published in English, will be included.

Research studies focusing on implementation of early warning systems and its impact on reducing burden of disease in low-income and middle-income countries and whose conclusions and discussion demonstrate transferable findings. All study designs will be considered including qualitative, quantitative and mixed-methods studies.

##### Exclusion criteria

Articles that are not in English will be excluded as the researcher will not assess them due to limited resources to translate research studies.

Research studies not focusing on implementation of early warning systems and its impact on reducing burden of disease.

##### Grey literature

Grey literature, including contemporary local government/agencies and charity reports, will be incorporated into the review to limit publication bias and ensure that all pertinent literature is located, but will be handled separately in terms of inclusion and exclusion criteria. The review team will determine how to handle the evidence from the grey literature once it has been retrieved.

##### Methodological quality assessment

All studies will be rated for quality using type of study specific quality appraisal tools from the Joanna Briggs institute [15].

Methodological quality assessment. All studies will be rated for quality using type of study specific quality appraisal tools from the Joanna Briggs institute [15].

These findings will be applied to the design and development of a pathogen and contaminant management tool. This pathogen and pollutant management tool will enable healthcare providers in the selected communities to identify and recognize emerging infectious diseases post flooding. This tool will be piloted and adapted with healthcare staff in collaboration with WP3. These findings will be applied to the design and development of a pathogen and contaminant management tool. This pathogen and pollutant management tool will enable healthcare providers in the selected communities to identify and recognize emerging infectious diseases post flooding. This tool will be piloted and adapted with healthcare staff in collaboration with WP3 and vital information can be transmitted via EWS to remote and isolated communities, facilitating the supply chain for prevention measures and treatment plans when flooding occurs. Finally, the piloted pathogen and pollutant management tool will be evaluated as part of WP4 Realist Evaluation and SROI. The systematic review protocol is registered on Prospero [16]

### Work package 2: Early Warning System Intervention (EWS) (0-48 months)

In WP2, will design, develop and evaluate the performance of an EWS and will link data on the two (rural and urban) healthcare and environmental contexts and integrate EWS for change, clarify how these will be delivered for vulnerable communities, and test and adapt the EWS.

#### Aim

To develop an integrated approach to monitoring and predicting EWEs such as flooding, prediction of the disease outbreak, and identification of the associated health risk for vulnerable communities in KZN province. This will improve the preparedness/response of the healthcare organisations/disaster management organisations and reduce healthcare burden. The EWS based on Artificial Intelligence (AI) and sensor technology for four vulnerable communities will be **developed for implementation by WP3** in two districts: eThekwini and Ugu districts of KZN province. An AI based solution will give vulnerable communities, health officials and policy makers early warning/alerts that they can use to make informed decisions, thereby limiting the impact of EWEs.

#### Objectives

**Objective 2.1**: To monitor, classify and predict high resolution spatial-temporal rainfall (time, intensity, duration, amount and floods) based on environmental parameters such as temperature, pressure, humidity, wind speed for vulnerable communities in KZN province using low-cost sensors, mobile and fixed sensors and AI.

**Objective 2.2**: To understand the impact of flooding (utilizing time series analysis of the historical and real time climate and disease outbreak data) on vulnerable communities and identify the risks of disease outbreak.

**Objective 2.3:** To develop and implement an EWS comprising of a dashboard and free mobile app, for alert/warning providing location-specific information for vulnerable communities, community leaders, disaster management, and health care organisations improving the preparedness and response.

##### Tasks

**Task 2.1 AI enabled EWE predictions (0-18 months):** To deploy low-cost energy efficient, open source integrated, portable hardware platform and sensor network (communities/locations identified in WP1) for monitoring climate indicators (e.g. temperature, humidity, atmospheric pressure, and wind speed) and develop an algorithm based on Artificial Intelligence for the prediction of EWEs such as flood and heavy rain.

**Task 2.2 AI enabled Disease outbreak predictions (0-24 months):** To design and develop an algorithm for the prediction of EWE-driven disease outbreak and associated risks by performing time series analysis of the historical EWEs and patient level data, and EWE prediction indicators (Task 2.1). This task will incorporate the evidence generated in **WP1 task 1.2** on the impact of flooding and early disease detection.

**Task 2.3 Design and development of an EWS (12-42 months):** To design, develop and launch a dashboard for a command-and-control centre to enable EWE and disease outbreak alert/warning for selected vulnerable communities and establish communication with healthcare organisations (input to WP3 for resource management), vulnerable communities, community leaders and disaster management organisations based on a cascaded structure model.

**Task 2.4 Test, verify and refine (24-48 months):** To emulate the EWE such as flooding in the selected vulnerable communities and test the prediction of disease outbreak and associated risks for verification and performance evaluation of alert/warning system. **WP4** evaluation will feedback data from consultation with communities for fine-tuning the system.

**Joint IP arising:** New climatic and health datasets linking disease outbreaks and EWE data. Applicants will remain compliant with the NIHR approach for IP sharing.

##### Method

Combined with (1) historical and real time meteorological data from the South African Weather Service (SAWS) along with sensors e.g., flood and high rainfall (2) emerging diseases outbreak data, e.g., diarrhoea, cholera, in this WP linked with evidence from **WP1 task 1.2**, the aim to exploit AI and sensing technology to identify the risk of disease outbreak due to flooding (e.g., disease outbreak, water contamination); examine the correlation between environmental indicators and disease outbreak and discover routes to improve the response of the disaster management and health care organisations through proactive EWS. The team will deploy sensing network (e.g., environmental sensors, weather stations and drones) in Isipingo, Merebank and Umnini (eThekwini) and Umzumbe (Ugu) across KZN (sites/communities identified in WP1) to monitor the environment in real-time (Task 2.1). The data from the sensing network will be collected via a central controller and transmitted to the cloud server (computers with data storage capabilities at UKZN) via the internet.

###### AI prediction for EWE

The University of KwaZulu-Natal (UKZN) will collaborate with University of the West of Scotland (UWS) to complement the existing computing capabilities and deploy a sensing network in Isipingo, Merebank and Umnini (eThekwini) and Umzumbe (Ugu) (sites/communities identified in WP1) to monitor the environment in real-time (Task 2.1). Data from the sensors and deployed drones to survey affected areas will be collected through a central controller and transmitted to the cloud server via the internet. Weather modelling is based on wide-area measurement and prediction augmented by ground-truth rain gauge measurements. The limitation of these techniques is the assumption of grid area average and large area weather fronts on the scale of the spatial resolution of the system used. The challenge in rain prediction is due to the complex interaction of weather parameters which include temperature, atmospheric pressure, humidity, wind speed and direction with the local topology. To minimise the impact of flooding, long-lead advanced warning with location-specific prediction is important to residents, the authorities and community organisations. Unlike most weather prediction systems, the developed prediction technique uses a simplified, cheaper sensor cluster measuring only temperature, humidity, atmospheric pressure, and wind speed to predict extreme weather conditions. This makes it cost-effective for expansion into other areas/communities.

###### Predicting disease outbreak using AI approaches

UKZN will collaborate with health care operators, the disaster management department for eThekwini and Ugu districts along with government portals to access the patient level symptomatic data and historical meteorological data. The healthcare data with timelines will be used to develop an AI disease outbreak predictive model for healthcare demands and the emergence of various conditions/symptoms in the selected vulnerable communities (Task 2.2). AI algorithms and predictive models developed by the University of the West of Scotland (UWS) team will be integrated with the EWS to predict flooding and heavy rain along with disease outbreak including risk identification for community-specific pre- and post-extreme event demands supporting the operations of the healthcare and disaster management organisations (input to **WP3** for resource management at healthcare providers). The model will account for the vector symptoms due to environmental factors and short to medium-term effects on healthcare sectors, professionals, medical supplies, and patients, especially those on treatment regimens for long-term conditions such as HIV and TB. The correlation between weather conditions and disease outbreaks will also be validated to benchmark the impact of climate change on vulnerable communities.

###### Alert and Warning System

We will deploy a command-and-control centre at UKZN with computer servers and uninterrupted electricity supplemented by backup generators when load shedding (a method to balance the demand and supply of electricity) occurs in conjunction with high-speed internet for hosting data securely, augmented with cloud-based storage, performing cloud computing, visualisation of the data and delivery of alert and warning in the event of heavy rains, flooding and disease outbreak (Task 2.3). We will design and develop a free-to-download mobile phone app with zoomable map and online dashboard (in at least two local languages) to deliver location-specific alerts/warning of flooding and disease outbreak in advance (7 days, 3 days and 1 day) for vulnerable communities. We will also create an interactive dashboard for the healthcare sector and disaster management organisations to exploit the demographic trends of weather conditions and disease outbreak risks. The mobile phone app and dashboard will provide real-time information on weather events to communities with location-specific status (**green, amber**, and **red**). With over 99.99% 3G coverage and 97.8% 4G coverage available throughout South Africa, it is expected that the population in these selected vulnerable communities already have access to high quality internet and smartphones [17]. However, we will adopt a robust approach to deliver the alert/warning in absence of smartphone or internet or related infrastructure. This will include, but is not limited to, advanced warning to community leaders and authorities and use of emergency sirens/SMS for communities at risk of immediate catastrophic adverse weather/flooding signifying risk to life.

##### Test Verification and performance

1. Test the accuracy of the rain (time of rain, rain intensity, rain amount and duration), flood and disease outbreak prediction system against measurements or required number of data observations. Establishment and testing of warning status protocol for different locations in the study area, e.g., verification of the grouping of the geographical areas into and **green, amber, red** alert/warning for flood and disease outbreak. This alert can be augmented with: **For the public** – avoid tap water or go to elevated spaces. **Community leaders** – guide public **Hospital** – check staff, resources etc
2. Tests of the early warning communication protocols which will reach the populations of eThekwini and Ugu Municipality districts (N= 265,767) and community responses in collaboration with **WP4** by examining the evaluation acceptability of EWS from the data returned from the 1520 respondents invited to complete the Social Return on Investment (SROI) questionnaire over the period of test time from the selected communities.

##### Benefits of EWS

###### Resilience in Health Services

Our approach will offer routes to improve the health care and disaster management organisations response, support preventive clinical management and control the infection through data analytics, computing technologies and community engagement in a considered case study. The health sector in KZN being largely publicly funded with limited resources, finding a disease outbreak risk tool will undoubtedly aid both medical staff and decision makers in better preparation and planning in a proactive manner. The health system would be better prepared in identifying high risk patients for prioritisation regarding early intervention and management.

###### Capacity Strengthening in LMIC

In LMICs, a small portion of the national budget is allocated to healthcare (e.g., less than 9% of the South African governments consolidated total expenditure). Therefore, in case of disease outbreak due to extreme climate conditions, supply of required medication etc. cannot be met. Our system’s data collection and analysis capability will act as an early-warning system, giving the government time to build treatment capacity. Capacity building also lies in the geographical spread of healthcare facilities across South Africa (scalability of the project). The project will also address resource limitations in South Africa such as better allocation of resources e.g., intensive care staff, supplies, drugs, etc.

###### Community Engagement

The community-based approach to the development of EWS will engender ownership, trust, and utilisation of the system in the community. The use of the existing Weather Department’s weather stations and augmentation with low-cost sensor clusters with cloud-based predictive computing reduces system failure risk and ensures application areas beyond the initially selected communities.

### Work package 3: Health system resilience and response to community health needs (0-48 months)

WP3, will develop a health system resilience and response to community health needs.

#### Aim

To ensure the health systems is resilient and able to respond to the health needs of communities at the time of an extreme weather event with a particular focus on flooding affecting the Ugu and eThekwini districts.

#### Objectives

**Objective 3.1 –** To assess the disease burden, health needs and community experiences at the time of an extreme weather event.

**Objective 3.2 –** To assess the resilience of the health system to respond in an extreme weather event.

**Objective 3.3 –** To train and mentor healthcare workers capacity to prepare for and adapt to increasing climate-sensitive disease risks and outbreaks ensuring a resilient health system and respond in EWEs.

In order to understand the disease burden, this WP will adopt a multipronged approach involving the health system and the community. This will be achieved through both secondary and primary data collection and analysis. For the secondary data analysis, data will be extracted on key disease outcome indicators from the KwaZulu-Natal District Health Information System. Amongst the data indicators considered will be incidence of diarrhoea in under five-year-old children, respiratory illness and schistosomiasis infections. The data will be extracted from 2017 till 2022 allowing for a trend analysis of the data from 2017 till 2022 with specific attention to periods related to adverse events in the province and chosen study sites. The knock-on effect on health service utilization and outcomes for other priority programmes (e.g. TB, HIV, COVID) will also be assessed. Primary data collection will entail a review of clinic and hospital outpatient registers and records to obtain details on demographic, and morbidity profiles for patients accessing healthcare services over the immediate period in 2022 after extreme climate events (see Table 1). In addition, this objective will take account of evidence gathered in **WP1 tasks** along with AI disease outbreak data from **WP2** with the aim to exploit AI and sensing technology to identify the risk of disease outbreak due to flooding.

**Table 1:**
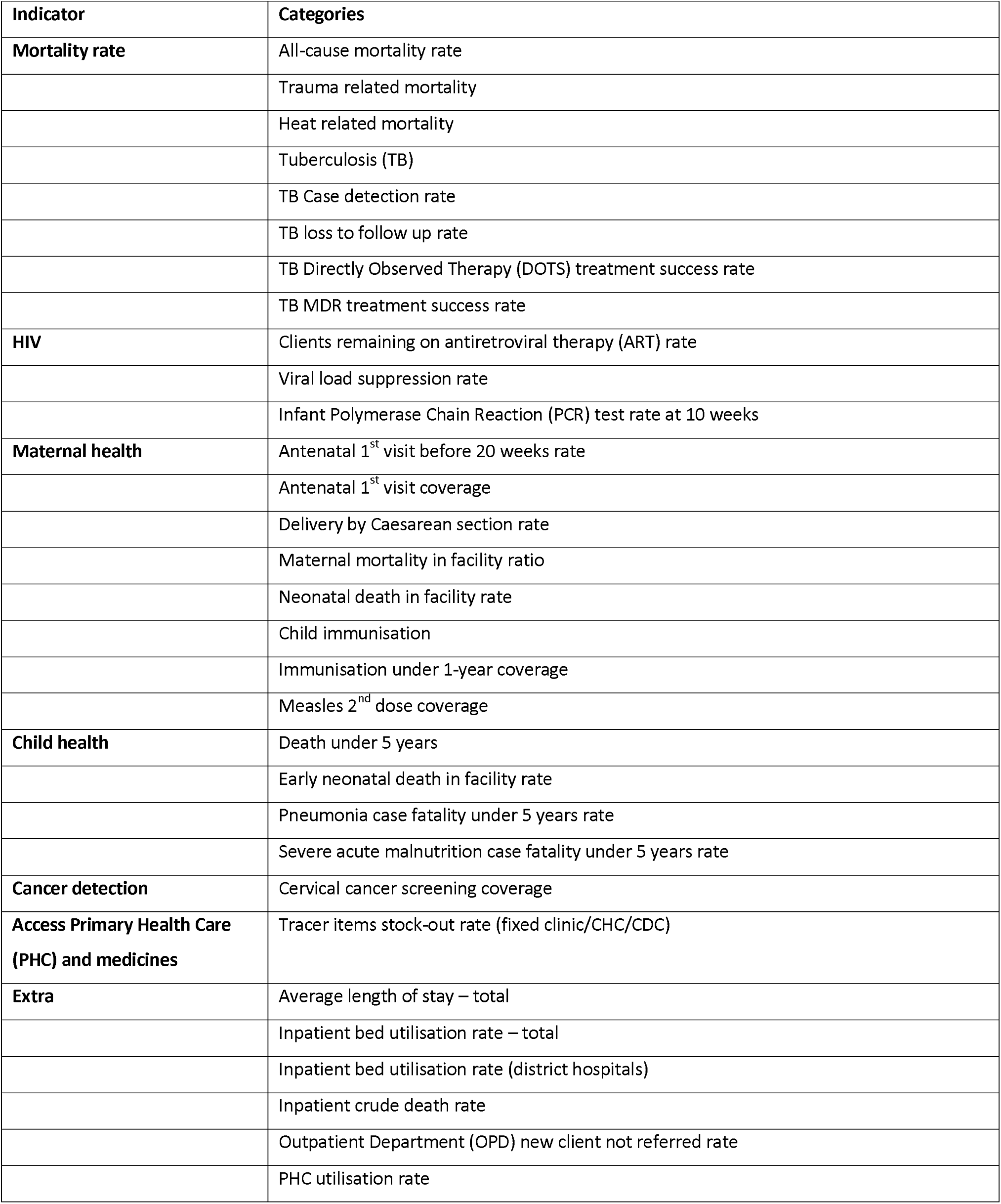
Variables on which information will be collected

| Indicator | Categories |
| --- | --- |
| **Mortality rate** | All-cause mortality rate |
|  | Trauma related mortality |
|  | Heat related mortality |
|  | Tuberculosis (TB) |
|  | TB Case detection rate |
|  | TB loss to follow up rate |
|  | TB Directly Observed Therapy (DOTS) treatment success rate |
|  | TB MDR treatment success rate |
| **HIV** | Clients remaining on antiretroviral therapy (ART) rate |
|  | Viral load suppression rate |
|  | Infant Polymerase Chain Reaction (PCR) test rate at 10 weeks |
| **Maternal health** | Antenatal 1<sup>st</sup> visit before 20 weeks rate |
|  | Antenatal 1<sup>st</sup> visit coverage |
|  | Delivery by Caesarean section rate |
|  | Maternal mortality in facility ratio |
|  | Neonatal death in facility rate |
|  | Child immunisation |
|  | Immunisation under 1-year coverage |
|  | Measles 2<sup>nd</sup> dose coverage |
| **Child health** | Death under 5 years |
|  | Early neonatal death in facility rate |
|  | Pneumonia case fatality under 5 years rate |
|  | Severe acute malnutrition case fatality under 5 years rate |
| **Cancer detection** | Cervical cancer screening coverage |
| **Access Primary Health Care (PHC) and medicines** | Tracer items stock-out rate (fixed clinic/CHC/CDC) |
| **Extra** | Average length of stay – total |
|  | Inpatient bed utilisation rate – total |
|  | Inpatient bed utilisation rate (district hospitals) |
|  | Inpatient crude death rate |
|  | Outpatient Department (OPD) new client not referred rate |
|  | PHC utilisation rate |

To assess community experiences, health symptoms and health seeking behaviour during adverse weather events, we will conduct a community survey informed by focus group discussions (see supplementary file 1). Two focus group discussions, with a minimum of eight and a maximum of twelve participants, will be held in each of the selected study sites. Thematic analysis of the focus group discussions will inform the development of a questionnaire that will be used for the community survey. The questionnaire will be piloted with a sample of twenty people and adaptations made to improve reliability of the questionnaire prior to implementation of the survey (see supplementary file 2). A cluster sampling approach using the WHO 30x7 method will be to determine the households that will participate in the survey. Specific sample size has been calculated based on population size using 95% confidence and an error of 0.05 (see Table 2).

**Table 2:** Sample size in the four regions

| Region | N | Sample size (n) |
| --- | --- | --- |
| **eThekwini** |  |  |
| Merebank | 40,000 | 380 |
| Umnini | 17,416 | 376 |
| Isipingo | 47,376 | 381 |
| **Ugu** |  |  |
| Umzumbe | 160,975 | 383 |
| **Total** | 265,767 | 1,520 (target invited sample) |

##### Objective 3.2

This objective will involve key informant interviews at provincial, district and facility level to establish the level of Leadership and Governance concerning climate resilience in the health system and a policy review (see supplementary file 3). Risk assessments informed by the WHO operational framework on building climate-resilient health systems [10] and the WHO guidance for climate-resilient and environmentally sustainable health care facilities [18] will be conducted in the chosen health districts and the health facilities in these districts. The risk assessments will focus on the components required to build climate resilient health systems including Leadership and Governance, Health work force capacity building, responsive health information systems and surveillance, service delivery and climate resilient technologies and infrastructure (see supplementary file 4). All primary health care facilities (n=8) and the respective hospitals (6) in the selected study sites will be reviewed to assess their climate resilience guided by the WHO guidance for climate-resilient and environmentally sustainable health care facilities. In addition, data will be accessed from Wentworth Hospital (eThekwini) Healthcare dashboard for resource management, the dashboard for disaster management along with the command-and-control centre at UKZN. The monitoring and reviewing of the dashboard data along with the evaluation of EWS in WP4 will inform the effectiveness and acceptability of the EWS for risk management and developing the resilience of health systems. Field workers with undergraduate training in health and safety will be trained by the research team and will pilot the pathogen and pollutant management tool designed and developed in WP1 which will enable healthcare providers to identify and recognise emerging infectious diseases post flooding.

This tool will be adapted and tested with field workers and healthcare staff to communicate the EWS to facilitate the supply chain for prevention measures and treatment plans when flooding occurs.

Finally, the tested pathogen and pollutant management tool will be evaluated as part of WP4 Realist Evaluation and SROI examining the effectiveness of the pathogen and pollutant management assessment tool to conduct the assessments.

##### Training

The training content of modules will be informed by the outcomes of the risk assessment and key informant interviews and will be co-created by subject and field experts drawn from Academia and the South African Department of Health, and through our links with the Regional Office of the WHO and Africa Centre for Disease Control and Prevention (Africa CDC). Modules will be developed and delivered over a 12-month period. Modules will focus on leadership skills, risk assessment and disaster management, using data to inform decision making and planning in the health service.

Training will consist of didactic teaching, group work, case studies and workplace-based exercises delivered through a combination of in-person and online sessions. Pre- and post-training evaluations will be conducted to assess changes in knowledge and practice. The training of healthcare workers will go beyond the facilities in the study sites and will be extended to the relevant healthcare workers in health facilities in all 11 health districts in KZN. An expected minimum of 200 healthcare workers will be trained. During the training the training team will identify training champions to be trained as Master trainers who in turn will cascade the training to the lower levels of staff within facilities including Community Health Workers (CHWs) in the Ward-Based primary health care Outreach Teams (WBOTs).

The training will be coupled with group and individual mentoring of facility managers and Master trainers within the health districts to ensure measures required for facilities and the health system to become resilient in EWEs are implemented. To ensure sustainability feedback meetings will be held quarterly in the first two years and then six monthly thereafter. (See Table 3 for logic model for objectives 3.2 and 3.3).

**Table 3:** Logic model for Objectives 3.2 and 3.3

| Objective | Activity | Output | Outcome |
| --- | --- | --- | --- |
| **1. Assess disease burden** | 1. Access and analysis of data from DHIS (2017 to 2022) | 1. Trend analysis of disease patterns during adverse events (2017-2022) | Overall picture of disease burden at the time of adverse events and service utilisation |
|  | 2. Primary data collection from health facilities for 2022 | 2. Disease burden at time of adverse event in 2022, health facility utilisation |  |
|  | 3. Service utilisation information for TB, HIV, COVID programmes in facilities | 3. Comparisons of service utilisation of priority programmes |  |
| **2. To assess health needs and community experiences at the time of an extreme weather event** | 1. Focus group discussions | 1. Qualitative view on community's health needs, service use and disease during extreme event | Report on the experience of communities, disease and service utilisation |
|  | 2. Pilot study of questionnaire | 2. Quantitative survey findings of disease, health needs and service use |  |
|  | 3. Questionnaire development |  |  |
|  | 4. Community survey |  |  |
| **Objective: To assess the resilience of the health system to respond in an extreme weather event.**<br>**1. Leadership and Governance** | 1. Key informant interviews with senior management in Department of Health | 1. Identify climate and health focal points in the department of health and district | Climate resilient policy at provincial, district and facility level |
|  | 2. Desktop Policy and legislative review | 2. Establish links between climate sensitive programmes and relevant departments outside of health required to respond to adverse events | Identify gaps for interventions |
| **2. Health work force and facility assessment** | 1. Survey of health workforce knowledge awareness and practice | 3. Percentage health workforce with knowledge and training |  |
|  |  | Percentage of facilities which are climate resilient |  |
|  | 2. Risk assessment of facilities to establish climate resilience in terms of WHO Building blocks |  |  |
|  | 3. Investigate EWS dashboard effectiveness and acceptability |  |  |
|  | 4. Pilot and adapt pathogen and pollutant management tool |  |  |
| **Objective 3: To train and mentor healthcare workers on how to ensure a resilient health system and respond in extreme weather events** | 1. Training programme (Policy development, risk assessment, disaster management, emergency preparedness) | 1. Training of 200 HCWs Pre-post training outcomes 2. Evaluation of mentoring | Build capacity in health workforce- evidenced by policies and practice |
|  | 2. Mentorship –group and individual |  |  |
|  | 3. Quarterly support meetings |  |  |

## Data analysis

The data analysis will be informed by the data collected. In summary, descriptive and inferential statistical analysis will be conducted on questionnaire data to gain a clear understanding about pathogen leading to disease outbreaks and risk management strategies development for health systems management during flooding. In terms of descriptive analysis, the functioning of health system components, frequencies of patients presenting with disease (diarrhoea, respiratory disease, schistosomiasis) and incidence rate ratio of water borne diseases amongst others will be reported on. Poisson regression analysis will be used to examine association between health system components and number of patients presenting with water borne disease. Generalized structural equation models (GSEM) with a Poisson response variable will be constructed to test direct and indirect effects of health system building blocks on incidence of water borne diseases following EWEs. Focus group data will be analysed via thematic analysis by means of qualitative analysis software, NVivo [19].

### Timeline and deliverables

See Gantt Chart (see supplementary file 5).

### Work package 4: Economic analysis including social return on investment (SROI)

#### Aim

The WEATHER study will examine the mechanisms of action and estimate the social costs and benefits of the EWS. A novel approach that combines Realist Evaluation (RE) and Social Return on Investment (SROI) in a longitudinal, mixed methods process, outcomes, and economic evaluation will be used. This involves the economic data collection happening alongside the RE to inform future sustainable and adaptable implementation via a continuous feedback loop established between work packages.

#### Objectives

SROI analysis will be used to value the change that the EWS makes to the outcomes that matter to people and the associated social value generated when people’s lives are improved owing to the successful combination of resources, input and processes. Community engagement activity, longitudinal focus groups and interviews (at 3 time points) will provide data for the evaluation and SROI, which have overlapping data collection needs. Focus groups and interviews with key informants within the healthcare system will be used to map both rural and urban healthcare contexts with attention focused to the role of local health systems, service provision and what might impact on achieving the desired outcomes related to minimising the impact of EWEs on health systems. Context mapping will contribute to establishing an initial programme theory, which will be refined with community engagement input to develop hypothesised Context Mechanism Outcome (CMO) configurations.

##### Focus groups with communities in South Africa

Focus groups consisting of 6-8 people approximately will be conducted in each of the four communities (Umgababa, Merebank, Isipingo and Port Shepstone) and 5-10 in-depth interviews with key informants (Community Health Workers (CHW)’s, health managers) will be held to determine likely mechanisms of change for the desired outcomes. This will focus on human resources, health facilities and experiences of EWEs. Outcome assessment at baseline and post intervention: WHO healthcare resilience; disease burden related to EWEs. In addition, should flooding occur due to EWEs during the lifetime of the project (0-48month) the team will take the opportunity to undertake a natural experiment which would involve collecting data through the WEATHER project under real conditions of flooding.

##### Focus groups and interviews with people implementing the intervention

Focus groups (two per community) and 5-10 interviews with those implementing the intervention to explore barriers and facilitators to implementing the EWS, to feedback to Work Package 2 for refining/optimising the intervention (end of year 2; repeated end of year 3).

##### Analysis of process and outcomes data

This task will include the analysis of the pathogen and pollutant management tool. The process evaluation will explore the outcomes (noted above) within the environmental, healthcare and population contexts in which they are achieved to establish a refined programme theory that will best inform future sustainable implementation in other settings [20], [21], [22].

Outcome data will be analysed to determine pre- and post-EWS variance as well as WHO resilience data to establish significance. The qualitative data will be recorded, and verbatim transcripts will be uploaded into NVivo [19].

A thematic analysis will seek to explore key questions related to intervention implementation and experience of the change in services. Implementation issues include exploring how the early warning system is used, applying early warning to healthcare delivery/preparedness, communicating public health messages and flood warnings to communities. This will be analysed alongside data on how this is experienced by those living in affected communities with a view to contextualising both the ‘soft’ and measurable outcomes and understanding how the system might be improved still further in the future. Matrix coding queries will be used to explore patterns across settings and attention will be paid to identifying and explaining disconfirming cases.

An overarching SROI will be conducted to estimate social cost benefit analysis taking a rigorous mixed method evaluation linked with study sample [23], [24]. The population of KZN province is 11, 065,240 million people making it the second most populous province in SA. There are 2.4% of the population or 265,767 individuals residing between the Umgababa, Merebank, Isipingo and Port Shepstone communities. Utilising these population statistics, the following equation was used to calculate the minimum sample size necessary to be representative of the four communities with a margin of error of +/- 5% with a confidence level of 95% with a standard deviation of .5 and population proportion of 25%. Applying the sample size equation and calculation of required minimum sample size indicates a sample size of N=384 respondents needed for the study to be representative of individuals residing in the four selected communities affected by flooding in KZN province, SA. The study will aim to attain at least a response rate of 25% and therefore will invite 1,536 participants to take part in the survey to achieve the required sample size based on the expected response rate.

##### Survey data

Quantitative data will be collected by means of SROI questionnaires from Umgababa, Merebank, Isipingo and Port Shepstone communities (see supplementary files 6 and 7). The SROI questionnaires will include individual outcome measures SWEMWBS (Shortened Warwick Edinburgh Mental and Emotional Wellbeing Scale) along with demographic individual data e.g., age, employment status, community settlement, income to examine measures of deprivation and explore if EWS impact on health inequities and inequalities in the two vulnerable KZN districts. Economic evaluation will include sensitivity analysis, discounting, opportunity costs, and Social Value Ratio (SVR).

## Discussion

Based on the study findings, recommendations will be made to the Department of Health with respect to an early warning system and health systems resilience. All information obtained will be anonymised, password-protected and accessible only to the investigators and stored with appropriate security.

## Declarations

### Ethics approval and consent to participate

#### Institutional ethical review board

This research protocol will be submitted to the Biomedical Research and Ethics Committee (BREC) at University of KwaZulu-Natal for expedited ethics approval, prior to the commencement of the study. Ethical approval will also be sought from the University of the West of Scotland as well as Royal College of Surgeons Ireland

#### Gatekeeper permission

The project will be registered with the National Health Research Ethics Committee and permission will be obtained from the KwaZulu-Natal Department of Health for the commencement of the study. Further gatekeeper permission will be obtained from all health facilities included in the study.

#### Informed consent

Every participant will sign an essential informed consent. All study participants will be guaranteed of confidentiality. In the questionnaire there will be no personal information recorded excluding demographic data. Study participants will be assured that participation is voluntary and that they can withdraw from the study at any stage should they wish to do so. The participants will be notified that there will be no financial incentive or direct benefits in participating in the research. Research results will be made available to participants on request to ensure transparency.

## Consent for publication

Not applicable.

## Availability of data and materials

Data sharing is not applicable to this article as no datasets were generated or analysed during the current study.

## Detailed work plan

The Gantt chart can be seen in supplementary file 5.

## Data Availability

All data produced in the present study are available upon reasonable request to the authors

## Acknowledgements

We would like to thank South Durban Community Environmental Alliance (SDCEA) facilitating all CEI activity and the community members from Umnini, Merebank, Isipingo (eThekwini) and Ugu in KwaZulu-Natal province, ZA for sharing their lived experiences during EWEs with the research team in developing this proposal.

## Authors’ contributions

**Conceptualization:** ML, SN, FH, MI, FHM, MZS, MG, DN, SV, OM, ND, CM.

**Funding acquisition:** ML and SN

Investigation: ML, SN, MI, LHS, SV, OM, MG, NNR, FN, FHM, MZS, ND, CM and DN. Methodology: ML, LHS, SN, SV, OM, MG, NNR, FN, FHM, MZS, ND, CM and DN.

Project administration: ML, SN, NNR and LHS

Supervision: ML, LHS, SN, SV, OM, MG, NNR, FN, FH, MZS, ND, CM and DN.

Visualisation: NNR

Writing – original draft: ML, FH, MI, SN, SV, OM, MG, NNR, FHM, MZS, and ND

Writing – review and editing: ML, FH, LHS, SN, SV, OM, MG, NNR, FN, FHM, MZS, and DN.

