## Supplementary File 1 for "Warning system for Extreme weather events, Awareness Technology for Healthcare, Equitable delivery, and Resilience (WEATHER) Project: A mixed methods research study protocol"

**KEY INFORMANT INTERVIEWS: FACILITY MANAGERS / OR INFRASTRUCTURE MAINTENANCE**

**Section A: Demographic data section**

Age

Gender Male Female

Profession

Position

Experience in health sector

**Open ended questions**

1. Do you think that climate change can impact the health of South Africans? If no, explore why. If yes, who do you think will be most affected and in what way?

 ______________________________________________________________________________
 ______________________________________________________________________________
 ______________________________________________________________________________
 ______________________________________________________________________________
 ______________________________________________________________________________
 ______________________________________________________________________________
 ____________________________________

2. Are you aware of the health complications from extreme weather events as a result of climate change in this province? Can you describe some of the complications that you have seen?

 ______________________________________________________________________________
 ______________________________________________________________________________
 ______________________________________________________________________________
 ______________________________________________________________________________
 ______________________________________________________________________________
 ______________________________________________________________________________
 _____________________________________

3. What climate change and health policies are in place, or should be in place? Does our health department have a policy that you are aware or a strategy in place to deal with climate change and health related complications?
 ______________________________________________________________________________
 ______________________________________________________________________________
 ______________________________________________________________________________
 ______________________________________________________________________________
 ______________________________________________________________________________
 _______________________________

4. How can climate change and health be aligned with Department of Health priorities? How do you think climate change and health intersects with National Health Insurance (NHI) programme?

5. What are the current strengths of the health facilities in dealing with extreme weather events as a result of climate change? How do you think this contributes to a resilient health system locally and provincially?

6. What are the current weaknesses of the health facilities in dealing with extreme weather events as a result of climate change? How do you think this contributes to a resilient health system locally and provincially?

7. How can climate change and health information be utilised so that it can help inform management, planning and governance? What do you think can be done to address restorative justice for communities that have been exposed to extreme weather events as a result of climate change?

8. Who do you suggest should be the stakeholders at the different levels that we should bring around a table to discuss a climate change and health plan?

________________________________________________________________________________
________________________________________________________________________________
________________________________________________________________________________
________________________________________________________________________________
________________________________________________________________________________
______________________________
