## Supplementary File 2 for "Warning system for Extreme weather events, Awareness Technology for Healthcare, Equitable delivery, and Resilience (WEATHER) Project: A mixed methods research study protocol"

**Sample questionnaire: START OF STUDY QUESTIONNAIRE FOR PARTICIPANTS**

**PLEASE NOTE:** Your contact details and answers will be treated as strictly confidential, and all data will be anonymised. Only the UKZN & UWS research teams will have access to the information you provide.

**SECTION 1**

**Q1. Name** ______________________________________________

**Q2. Age** ____________

**Q3. Gender**

☐ Male ☐ Female ☐ Prefer not to say

**Q4. Ethnic origin**

☐ Indian/Asian ☐ Black African ☐ Coloured ☐ Mixed ☐ White

☐ Other (please specify) ______________________________

**Q5. Community**: __________________________________________

**Q6. Occupation**: __________________________________________

**Q7. Email**: ______________________________________________

**Q8.**

Please rank the following reasons for joining this research in order of relevance you to, from 1 to 8 (1 = most relevant to you, 8 = least relevant to you).

☐ Explore/ return to crafting

☐ Improve mental health & wellbeing

☐ Improve physical health & wellbeing

☐ Learn something new

☐ Meet new people

☐ Reduce fatigue

☐ Reduce stress and/ or anxiety

☐ Spend time in my community

**Q9. In addition to the above, is there anything else you hope to experience from participating in the predictive early warning system WEATHER project?**

____________________________________________________________________________

____________________________________________________________________________

**SECTION 2**

**Q10. For each statement below, please tick the box that best describes your experience over the past 2 WEEKS.**

| STATEMENTS | None of the time | Rarely | Some of the time | Often | All of the time |
|---|---|---|---|---|---|
| I’ve been feeling optimistic about the future | | | | | |
| I’ve been feeling useful | | | | | |
| I’ve been feeling relaxed | | | | | |
| I’ve been dealing with problems well | | | | | |
| I’ve been thinking clearly | | | | | |
| I’ve been feeling close to other people | | | | | |
| I’ve been able to make up my own mind about things. | | | | | |

**Q11. Please indicate how often you’ve used the following health services in the last 8 WEEKS.**

| In the last 8 weeks, how many appointments have you had with the following local health staff or services? | Number of online appointments | Number of face-to-face appointments |
|---|---|---|
| GP | | |
| Community nurse | | |
| Hospital | | |
| Mental health nurse | | |
| Community Health Worker | | |
| Physiotherapist | | |
| Psychiatrist | | |
| Psychologist | | |
| Social worker | | |
| Other (please state)___________________________________ | | |

**SECTION 3: How you're feeling now**

**Q12. Under each heading, please tick the ONE box that best describes your health TODAY.**

| **Mobility** | **Tick one option below** |
|---|---|
| I have no problems in walking about | |
| I have slight problems in walking about | |
| I have moderate problems in walking about | |
| I have severe problems in walking about | |
| I am unable to walk about | |
| **Self-Care** | **Tick one option below** |
| I have no problems washing or dressing myself | |
| I have slight problems washing or dressing myself | |
| I have moderate problems washing or dressing myself | |
| I have severe problems washing or dressing myself | |
| I am unable to wash or dress myself | |
| **Usual Activities** (e.g., work, study, housework, leisure activity) | **Tick one option below** |
| I have no problems doing my usual activities | |
| I have slight problems doing my usual activities | |
| I have moderate problems doing my usual activities | |
| I have severe problems doing my usual activities | |
| I am unable to do my usual activities | |
| **Pain / Discomfort** | **Tick one option below** |
| I have no pain or discomfort | |
| I have slight pain or discomfort | |
| I have moderate pain or discomfort | |
| I have severe pain or discomfort | |
| I have extreme pain or discomfort | |
| **Anxiety / Depression** | **Tick one option below** |
| I am not anxious or depressed | |
| I am slightly anxious or depressed | |
| I am moderately anxious or depressed | |
| I am severely anxious or depressed | |
| I am extremely anxious or depressed | |

**Q13. Please indicate which statements best describe your overall quality of life at the moment by placing a tick in ONE box for each of the five groups below.**

| **Feeling settled and secure** | **Tick one option below** |
|---|---|
| I am able to feel settled and secure in **all** areas of my life | |
| I am able to feel settled and secure in **many** areas of my life | |
| I am able to feel settled and secure in **a few** areas of my life | |
| I am **unable** to feel settled and secure in **any** areas of my life | |
| **Love, friendship and support** | **Tick one option below** |
| I can have **a lot** of love, friendship and support | |
| I can have **quite a lot** of love, friendship and support | |
| I can have **a little** love, friendship and support | |
| I **cannot** have **any** love, friendship and support | |
| **Being independent** | **Tick one option below** |
| I am able to be **completely** independent | |
| I am able to be independent in **many** things | |
| I am able to be independent in **a few** things | |
| I am **unable** to be at all independent | |
| **Achievement and progress** | **Tick one option below** |
| I can achieve and progress in **all** aspects of my life | |
| I can achieve and progress in **many** aspects of my life | |
| I can achieve and progress in **a few** aspects of my life | |
| I **cannot** achieve and progress in **any** aspects of my life | |
| **Enjoyment and pleasure** | **Tick one option below** |
| I can have **a lot** of enjoyment and pleasure | |
| I can have **quite a lot** of enjoyment and pleasure | |
| I can have **a little** enjoyment and pleasure | |
| I **cannot** have **any** enjoyment and pleasure | |

**Q14. For each statement below, please tick the box that best describes your experience NOW.**

| STATEMENTS | Strongly Disagree | Disagree | Neutral | Agree | Strongly Agree |
|---|---|---|---|---|---|
| I am content with my friendships and relationships | | | | | |
| I have enough people I feel comfortable asking for help at any time | | | | | |
| My relationships are as satisfying as I would want them to be | | | | | |

**Q15. For each statement below, please tick the box that best describes your experience NOW.**

| STATEMENTS | Not at all true | Hardly true | Moderately true | Exactly true |
|---|---|---|---|---|
| I can always manage to solve difficult problems if I try hard enough | | | | |
| If someone opposes me, I can find the means and ways to get what I want | | | | |
| It is easy for me to stick to my aims and accomplish my goals | | | | |
| I am confident that I could deal efficiently with unexpected events | | | | |
| Thanks to my resourcefulness, I know how to handle unforeseen situations | | | | |
| I can solve most problems if I invest the necessary effort | | | | |
| I can remain calm when facing difficulties because I can rely on my coping abilities | | | | |
| When I am confronted with a problem, I can usually find several solutions | | | | |
| If I am in trouble, I can usually think of a solution | | | | |
| I can usually handle whatever comes my way | | | | |

**SECTION 4: You and your local community**

**Q16. Please tell us how often you agree with each of the following statements by putting a tick in the relevant box.**

| STATEMENTS | Completely disagree | Strongly disagree | Disagree | Neither agree or disagree | Agree | Strongly agree | Completely agreed |
|---|---|---|---|---|---|---|---|
| I always find beauty in my local community | | | | | | | |
| I always treat community with respect | | | | | | | |
| Being in community makes me very happy | | | | | | | |
| Spending time in community is very important to me | | | | | | | |
| I find being in community really amazing | | | | | | | |
| I feel part of the community | | | | | | | |

**Thank you for completing this questionnaire.**

**Your participation in this study is much appreciated.**

Please note that your contact details and answers will be treated as strictly confidential, and all data will be anonymised. Only the UKZN and UWS research teams will have access to the information you provide.
