## Supplementary File 3 for "Warning system for Extreme weather events, Awareness Technology for Healthcare, Equitable delivery, and Resilience (WEATHER) Project: A mixed methods research study protocol"

**Focus group/stakeholder engagement questions for Social Return on Investment (SROI) analysis exploring the social value generated by the Weather project Early Warning System.**

**Involve stakeholders:**

Inform what gets measured and how this is measured and valued by involving stakeholders. Stakeholders are those people or organisations that experience change as a result of the activity and they will be best placed to describe the change. This principle means that stakeholders need to be identified and then involved in consultation throughout the analysis, in order that the value, and the way that it is measured, is informed by those affected by or who affect the activity. This phase is about defining outcomes and in informing the relative importance of the outcomes.

**Understand what changes:** Articulate how change is created and evaluate this through evidence gathered, recognising positive and negative changes as well as those that are intended and unintended. Value is created for or by different stakeholders as a result of different types of change; changes that the stakeholders intend and do not intend, as well as changes that are positive and negative. This principle requires the theory of how these changes are created to be stated and supported by evidence. These changes are the outcomes of the activity, made possible by the contributions of stakeholders, and often thought of as social, economic or environmental outcomes. It is these outcomes that should be measured in order to provide evidence that the change has taken place.

**Value the things that matter:** Use financial proxies in order that the value of the outcomes can be recognised. Many outcomes are not traded in markets and as a result, their value is not recognised. Financial proxies should be used in order to recognise the value of these outcomes and to give a voice to those excluded from markets but who are affected by activities. This will influence the existing balance of power between different stakeholders.

The **aim** of this initial phase of the SROI is to guide the co-design and co-production and the identification of inputs, outputs and health and wellbeing outcomes of the WEATHER project Early Warning System (EWS) and associated social value.

**Objectives**

1. To determine the range of stakeholder expectations and desired outcomes from the Weather project EWS
2. To identify the wellbeing outcomes
3. To recognise the wide-ranging stakeholder expectations and requirements
4. To develop a systematic theory of change and accompanying outcomes framework

**Methods:**

The initial steps of the SROI will develop and make explicit the narrative of how and why a desired change (resilience, purpose/direction, satisfaction, connection, etc.) is expected to happen in a particular context (as a result of Weather project EWS). It maps or 'fills in' the middle bit between what a project does (its activities or interventions) and how these achieve project goals will be achieved.

The engagement of stakeholders leads to an understanding does this by first identifying the desired long-term goals and then working back from these to identify all the conditions (outcomes) that must be in place (and how these related to one another causally) for the goals to occur and mapping of the outcomes. This then provides the basis for identifying what type of activity will lead to the outcomes identified as preconditions for achieving the long-term goal.

**Questions for SROI at initial engagement and mapping of outcomes and questionnaire development.**

**Inputs**

- How are you involved in the Weather project EWS?
- What did you contribute to the activity (and how much)?

**Activities**

- What activity/activities did you think you will experience/have you experienced?

**Outcomes-** Outcomes are changes that stakeholders experience as a result of the activity. We will be referring to outcomes as 'changes' for the rest of this guide. There are lots of different ways in which people can experience change. The taxonomy in this guidance recognises five main types of change that people can experience. These are changes in:

a) Circumstance b) Behaviour c) Capacity d) Awareness e) Attitude

- What changes do you think you will experience/have you experienced?
- What do think will be different/has been different as a result of the WEATHER EWS project?
- Do you think there will be/there have been positive changes?
- Do you think there will be/there have been negative changes?
- Do you think anyone else will experience/has experienced any changes as a result of the WEATHER EWS project?

**Indicators**

- How would you let someone else know that you experienced any changes and what would you show them?
- What would it look like?
- Could you measure it?

**Deadweight**

- What would have happened to you if you didn't take part in the WEATHER EWS project?
- Would you have found something else later?

**Attribution**

- Who else provides something like this in your local area?
- Do you think that anyone else contributes to the experience/change?

**Displacement**

- Do you think that you will have to give up anything to take part in the WEATHER EWS project?
- Are you getting similar support from somewhere else?

**Duration**

- How long do you think the change will last?
- Imagine we are 2 years or 5 years from now, do you think you'll still be experiencing the change?

**Valuation**

- How important do you think this change is to you?
- Can you compare it to something else just as important to you?
- Can you put these changes in priority order of how important they are to you?
- Which are worth the most / the least to you?
- Which of these changes will make the biggest difference to you?

| Principle | Achieved | Prompt |
|---|---|---|
| **Inputs** | **Yes** □<br>**No** □ | How are you involved in the WEATHER EWS project?<br>What did you contribute to the activity (and how much)?<br>What activity/activities did you think you will experience/have experienced? |
| **Outcomes** | **Yes** □<br>**No** □ | What changes do you think you will experience/have experienced?<br>What do think will be/has been different as a result of the WEATHER EWS project?<br>Do you think there will be/have been positive changes/negative changes?<br>Do you think anyone else will experience any changes as a result of the WEATHER EWS project? |
| **Indicators** | **Yes** □<br>**No** □ | How would you let someone else know that you experienced any changes and what would you show them?<br>What would it look like?<br>Could you measure it? |
| **Deadweight** | **Yes** □<br>**No** □ | What would have happened to you if you didn't take part in the WEATHER EWS project?<br>Would you have found something else later? |
| **Attribution** | **Yes** □<br>**No** □ | Who else provides something like this in your local area?<br>Do you think that anyone else contributes to the experience/change? |
| **Displacement** | **Yes** □<br>**No** □ | Do you think that you will have to give up anything to take part in the WEATHER EWS project?<br>Are you getting similar support from somewhere else? |
| **Duration** | **Yes** □<br>**No** □ | How long do you think the change will last?<br>Imagine we are 2 years or 5 years from now, do you think you'll still be experiencing the change? |

| | | |
|---|---|---|
| **Valuation** | **Yes** □<br>**No** □ | How important do you think this change is to you?<br>Can you compare it to something else just as important to you?<br>Can you put these changes in a priority order of how important they are to you?<br>Which are worth the most / the least to you?<br>Which of these changes will make the biggest difference to you? |
