## Supplementary File 4 for "Warning system for Extreme weather events, Awareness Technology for Healthcare, Equitable delivery, and Resilience (WEATHER) Project: A mixed methods research study protocol"

**Health Facility assessment for climate resilience and environmental sustainability**

**Section A: Health facility information**

**1.1Health facility information:**

**1.1.1 Facility Name:_______________________________________________**

**1.1.2. Health District:** **1. eThekwini** **2. Ugu**

**1.1.4. Facility type:**

**1. Tertiary hospital**

**2. Regional hospital**

**3. District hospital**

**4. Community Health Clinic**

**5. Primary Health Clinic**

**Section B: Health workforce**

**1.2 Key informant information:**

1.2.1. Age

1.2.2. Gender Male Female

1.2.3 Job description

1.2.4. Years employed in current job:

1.2.5. Highest Qualification:___________________________

1.2.6. Any training on facility climate resilience yes no

1.2.7: if yes provide details____________________________________________________

| **Section B: Health workforce** | | | | |
|---|---|---|---|---|
| **Questions** | **Unavailable; not done** | **In progress; incomplete** | **Completed; achieved** | **Observations** |
| **Human Resources (Climate resilience)** | | | | |
| Assessment of potential workplace hazards that may arise in emergencies, and planning to address measures to reduce those hazards | | | | |
| Do health workers and local communities work together to promote a health care facility environment safe from climate related impacts? | | | | |
| Are there established systems for management of occupational safety and health in all health care facilities? | | | | |
| Have you identified minimum needs in terms of health care workers to ensure the operational sufficiency of every health care facility department, in case of climate related disaster or emergency? | | | | |
| Is there any established system of rapidly providing health workers (such as voluntary medical personnel) with necessary credentials in an emergency, in accordance with health care facility and health authority policies? | | | | |
| Are there multidisciplinary psychosocial support teams in place for staff, families of staff and patients (such as in emergency and disaster situations)? | | | | |
| Is there an established post-disaster employee recovery assistance programme according to staff needs? | | | | |
| Is there an early warning system in place to respond to adverse events? | | | | |
| Do you have a contingency plan for personnel transportation in place to respond to emergencies*? | | | | |
| Is there a disaster risk reduction plan in place for the health workforce to manage measures of prevention, preparation response and recovery from extreme air pollution? | | | | |
| Is there a contingency plan in place for evacuation during or following an extreme event*? | | | | |
| Are there established mutual aid and assistance agreements (such as transfer of patients, sharing of resources and supplies) with other sectors or institutions to have health support (including of health workforce) during response and recovery from an extreme weather event or disaster? | | | | |
| Are there clearly defined security measures in place for safe and efficient hospital evacuation*? | | | | |

| |
|---|
| Is the health workforce able to assess potential health impacts and facility loss associated with climate related hazards? |
| Is there sufficient emergency room surge capacity available to manage air pollution climate-related emergencies diseases? |
| **Human resources (environmental sustainability)** |
| Have you ensured protection of the health workforce in vulnerable situations through environmentally sustainable practices? |
| Have measures been implemented by health workforce to eliminate disease burden among vulnerable populations resulting from environmental hazards in health care facilities? |
| Are you identifying opportunities to improve work practices in environmentally friendly ways, and integrating initiatives? |
| Has the health workforce been trained to implement environmentally sustainable interventions for infection prevention and control; and combating antimicrobial resistance? |
| Do you ensure rapid clean-up and recovery from extreme weather events to avoid indoor air quality problems (such as mould growth associated with floods)*? |
| Do the health care facility staff and patients drink filtered tap water when safe*? |
| Are the health care facility staff trained to assess their water use for implementing potential savings measures? |
| Do health care facility staff monitor and assess water drips, leaks, and unnecessary flows in bathrooms, laundry facilities, kitchen, etc. for prompt repairs*? |
| **Health workforce capacity development (Climate resilience)** |
| Which prevention and education programmes are done by healthcare staff to reduce disease burden of adverse weather events? |
| Is the health workforce trained to address climate change risks to health through WASH, and chemical and energy related hazards? |
| Have measures been implemented to diminish disease burden of climate related hazards by increased health actions of staff and community through prevention and education programmes*? |
| Is the Health workforce taking part in community educational programmes to assist the local community in reducing air pollution related risks? |
| Is there an Established Emergency Operational Committee or hospital Incident Command Group for climate related air pollution? |
| Are there systems in place for monitoring diseases from climate related hazards including monitoring |

| |
|---|
| health outcomes to health care workers and vulnerable patients (such as the elderly, immobile, infants, critical care patients) in the face of air pollution climate related emergency or disaster? |
| Is there a plan in place for relocating hospital equipment, medicines and medical devices during floods or permanent relocation of equipment to higher floors in flood-prone areas*? |
| Have you improved staff capacity to provide safe and reliable infection prevention and control services, when a disaster or emergency crisis occurs? |
| Is the Health care facility staff trained to identify health threats made worse by air pollution climate related events, to reduce associated morbidities from respiratory and cardiovascular diseases, nutrient deficiency, mental health issues? |
| Have training and exercises been provided in areas of potential increased clinical demand following a climate related event or outbreak to ensure adequate staff capacity and competency*? |
| Is the Health workforce trained to detect posttraumatic stress disorder of long-term air pollution of the population and take prompt action? |
| Is the health workforce trained (including exercises, simulations) for early warning system, contingency plan, and disaster preparedness, response and recovery management to address climate change risks and to cope with any emergency from climate related disasters and outbreaks, epidemics and pandemics*? |
| Have training and exercises been provided in areas of potential increased clinical demand following a climate related event or outbreak to ensure adequate staff capacity and competency*? |
| Are health care facility staff, responsible for critical systems, trained in emergency preparedness and response and to communicate effectively in emergency situations? |
| Does the health workforce receive training and exercises for preparing, responding and recovering from extreme weather-related emergencies*? |
| Are facility staff trained in protecting their health and safety during an emergency situation? |
| Can the health workforce implement safe water management in weather-related emergencies and disasters, according to local conditions and disaster magnitude*? |
| Is the Health workforce receiving training and exercises on surveillance systems for air pollution climate related diseases? |

| |
|---|
| Is the health workforce trained to an appropriate standard to maintain the correct level of safety of water quality controls, supplies and alternative sources to the health care facility in both routine and emergency/disaster situations*? |
| Is a plan in place for water system supplies (such as chlorine, filters or other water treatment technology, rapid water testing kit), during an emergency and disaster response*? |
| Has the been an increase in health workforce knowledge on waste stream constituents and waste related health care hazards for better monitoring and control in climate related emergency situations* |
| Are the health workforce trained to an appropriate standard to maintain correct level of chemical safety, and safety of waste management systems of the health care facility in both routine and emergency/disaster situations? |
| Are the health workforce trained to an appropriate standard to maintain correct level of safety of electrical power supply and alternate source (such as generators) of the health care facility in both routine and emergency/disaster situations? |
| **Capacity Development (environmental sustainability)** |
| Is education and training provided to health care facility staff and the community on environmental factors that contribute to the burden of disease? |
| Is education and training provided to health care facility staff and the community on the relationship between public environmental health and disease prevention? |
| Is education and training provided to health care facility staff and the community on how to evaluate and select environmentally sustainable products and services*? |
| Is access provided to environmental information and training, including priority setting approaches and effective procurement*? |
| Is the health workforce trained in the management of chemicals and health care waste? |
| Are you developing and implementing awareness campaigns about chemicals of concern and established best practices for safe chemicals management within the health sector? |
| **Climate resilience - Communication and awareness raising** |
| Are there any opportunities for health workforce to learn and increase awareness about climate change, its impacts, and co-benefits of sustainable practices? |

| |
|---|
| Is there any ongoing awareness campaigns of health care facility staff, patients, visitors, and the community of risks to health from air pollution and effective health protection measures? |
| Do you receive updated emergency plans as new knowledge on air pollution related risks become available? |
| Is the health workforce aware of approaches to childhood development and social outcomes related to nutrition and avoidance of stunting and impaired neurological development due climate change impacts on water supply, food production, infectious diseases? |
| Is the health workforce engaged in community health programmes to improve community health during particular climate risks (such as home care for asthma to reduce health vulnerabilities during episodes of high air pollution or heat waves)*? |
| Is there scheduling of outdoor work for cooler parts of the day and reduce physical demands during hot days? |
| Are you considering indoor and outdoor temperatures when planning group activities in hot days or heat waves? |
| Do health care facility staff help organize and participate in community disaster planning committees*? |
| Are key messages for target audiences (such as patients, staff, public) drafted in preparation for the most likely extreme weather disaster scenarios*? |
| Are there updated emergency plans as new knowledge on climate risks become available? |
| Are there awareness campaigns about chemicals of concern and established best practices for safe chemicals management? |
| **Communication and awareness raising (environmental sustainability)** |
| Is there increased awareness about water conservation? |
| Are health care facility staff informed on managing safe wastewater to combat antimicrobial resistance? |
| Does the health workforce recycle all different types of non-hazardous waste (uncontaminated paper, plastic, glass, metal)? |
| Is there increased knowledge about the environmental impact of pharmaceuticals and their disposal? |
| Does the health workforce understand the benefits associated with correct equipment and systems operation to save energy*? |

| |
|---|
| Does the health workforce use stairs and ramps, whenever possible, to reduce elevator usage and promote physical activity? |

**Section C: water, sanitation and health care waste interventions**

**Section A: Demographic data section**

**1.2 Key informant information:**

1.2.1. Age

1.2.2. Gender Male Female

1.2.3 Job description

1.2.4. Years employed in current job:

1.2.5. Highest Qualification:____________________________

1.2.6. Any training on facility climate resilience yes no

1.2.7: if yes provide details______________________________________________________

| **Section B: Water, Sanitation and Health Care Waste Interventions (Climate Resilience)** | | | | |
|---|---|---|---|---|
| **Monitoring and assessment (Climate resilience)** | | | | |
| | **Unavailable; not done** | **In progress; incomplete** | **Completed; achieved** | **Observations** |
| Are you maintaining safety conditions and proper functioning of all elements of the water distribution system, including storage tanks, valves, pipes and connections, and water disinfection? | | | | |
| Do you check Water pipe connections regularly for signs of deterioration? | | | | |
| Have you developed a monitoring mechanism to verify compliance with national standards, including the operation and maintenance of water and sanitation facilities? | | | | |
| Is the Water quality supply monitored regularly for air pollution environmental pollutants? | | | | |
| Are you identifying current or historical air pollution related hazardous events known to pose significant health risks to water system? | | | | |
| Have you developed climate-resilient water safety plans? | | | | |
| Is the water supply monitored regularly during emergencies to ensure adequate access | | | | |

| |
|---|
| throughout the duration of the event, ensuring that protocols are in place to guide rationing if required? |
| Is there monitoring of sewer overflows to fix pumps in advance of flood seasons? |
| **Monitoring and assessment (Climate resilience)** |
| Do you measure where and how water is used, and areas of potential savings and reuse examined? |
| Are measures implemented to conserve and save water in staff training especially during the induction of new staff? |
| Is there surveillance of diseases related to insufficient quality water, sanitation? |
| Have you classified and assessed the types of waste issues and hazards to establish segregation collection? |
| Have you implemented and monitored a waste reduction programme including waste management training for all staff? |
| **Risk management (climate resilience)** |
| Have you developed a long-term drought management plan, including the identification of available alternative safe water sources*? |
| Does the health care facility conserve and manage water to reduce water usage? |
| Are water services affected by seasonality or climate change related weather extremes*? |
| Is the WASH climate risk management plan implemented? |
| Is there improved training and support to health workforce on how and when to deliver water messaging? |
| Is safe water storage available, avoiding mosquito breeding sites*? |
| Is water contaminated in the health care setting during storage, distribution and handling*? |

| |
|---|
| Does the kitchen have adequate supplies of clean potable water*? |
| Do water storage tanks have appropriate covers to prevent access or contamination? |
| Are there non-return valves on water supply pipes installed to prevent back flows*? |
| Is water storage in the health care facility sufficient to meet the needs of the facility in case of water contamination from adverse events? |
| Is storm water safely managed (avoiding standing water near the facility or affecting nearly (households)*? |
| Is the health care facility drinking water treated with a residual disinfectant to ensure microbial safety up to the point of consumption or use as the area is known for high levels of environmental pollutants? |
| Are water storage tanks not located in areas susceptible to drainage from pollution during adverse events, thus reducing the risk of contamination? |
| Are plastic water storage tanks supported and anchored to resist strong winds? |
| Is there natural floodwater infiltration in place to reduce risk of facility flooding? |
| Are there assessments and mapping of climate change risks to the sanitation infrastructure of health care facilities in place to identify where services could be disrupted from floods, water scarcity, landslides, sea-level rise? |
| Is there a planned schedule for emptying latrines in advance of flood seasons to avoid overflows? |
| Is there installation of sealed covers for septic tanks and non-return valves on pipes to prevent back flows? |
| Are there vents on sewers and septic tanks are above expected flood lines? |
| Are the waste issues resulting from environmental pollution |

| |
|---|
| from adverse events assessed to establish safe procedures and specialized treatment, when needed? |
| Is health care waste transport (including health care facility hazardous waste) properly managed in case of extreme weather events*? |
| **Risk Management (Environmental sustainability)** |
| Is there reinforced messaging about water use through signs and notices to promote saving? |
| Is there increased patient and visitor awareness about water conservation including signs and notices in patient rooms and visitor restrooms? |
| Is plastic bottled water eliminated where a drinking tap water is available*? |
| Are eating utensils are washed immediately after use (reducing water and energy)? |
| Is wastewater safely managed through use of on-site treatment (such as septic tank followed by drainage pit) or sent to a functioning sewer system*? |
| Is there an established recycling programme for all types of non-hazardous waste*? |
| Is there established segregation collection of different types of waste according to hazards*? |
| Have you phased out of incineration of medical waste: a variety of non-burn technologies are available to safely disinfect, neutralize or contain waste (such as autoclaving)? |
| Does the waste disposal system include separate bins for potentially infectious waste, sharps, chemicals, pharmaceuticals, non-hazardous wastes? |
| Do cleaning products that contain hazardous chemicals such as those found in some soaps, disinfectants and pesticides are clearly labelled following the |

| |
|---|
| Globally Harmonized Classification System? |
| Is there reduced use of mercury-containing medical devices and measures in place to manage mercury spills and mercury-contaminated wastes*? |
| Is there mproved packaging, labelling and identification of chemical waste in separate chemical-resistant containers (i.e. not mixing hazardous chemical wastes of different types)*? |
| Is there improved packaging and identification of low-level radioactive waste that may be collected in containers clearly labelled with the international radioactive symbol and the words “radioactive waste”*? |
| Is there improved packaging, identification and storage of radioactive waste in containers that prevent dispersion of radiation (storage behind lead shielding)*? |
| **Health and safety regulation (climate resilience)** |
| Is the water of appropriate quality supplied for medical activities as well as for vulnerable patients (certain procedures should meet strict criteria and additional treatment or source, concerning microbial and chemical contaminants, including cyanobacterial toxins, and chlorine and aluminium which are commonly used in drinking water treatment)? |
| Are you collaborating with public health management or other responsible sector to reduce vector breeding sites from open mining holes on facility property and surrounding areas? |
| Is the Health care facility able to provide clean water for patients and the health workforce in the face of air pollution related disasters? |

| |
|---|
| Is there a readily available disaster response and recovery plan for the water system with adequate supplies (such as chlorine, filters or other water treatment technology, rapid water testing kit) given that this area is prone to adverse weather events? |
| Are the sanitation technologies designed to be more resistant during adverse events? |
| Is there rainwater harvesting (with safe storage) installed, in places where rainfall is sufficient and regular or when possible to collect, and regularly inspected for damage? |
| Is there a possibility that health care facility wastewater disposal contaminates local serviceable drinking water *? |
| Is a long term water collection system in place to ensure water access during extreme climate events (such as capturing rain during the monsoon season and storing water in tanks for use during the dry season)? |
| Have you ensured effective and timely delivery of safe water during emergencies over the short- and long-term*? |
| Is there improved storage areas for storing extra waste generated through higher demands on health care facilities (such as in outbreaks or impacts from climate related events)*? |
| Are waste pits built to withstand climate events and emergencies*? |
| Is the health care facility waste disposal safe during climate-related emergencies or disasters? |
| **Health and safety regulation (environmental sustainability)** |
| Have you established and informed staff on the water conservation policy? |
| Is harvested rainwater or grey water safely used to flush toilets, clean outdoor pavement areas, water plants when possible? |

| |
|---|
| Are monitoring systems in place for early detection and control of health care associated infections? |
| Are there hand hygiene facilities (water and soap and alcohol-based hand rub) available at points of care and before health care facility entry during outbreaks, epidemics and pandemics? |
| Are there hand hygiene facilities (water and soap and alcohol-based hand rub) available within five metres of all toilets? |
| Does the health care facility safely dispose of hazardous wastewater and liquid waste into the sanitation system through pre-treatment (such as oils and fats, corrosive waste and other wastes, depending on the level of concentration)*? |
| Does the health care facility safety dispose of hazardous wastewater and liquid waste that may be infectious*? |
| Do you facilitate the use of safer alternatives and sound management of health care waste, drawing on relevant guidance from WHO and others, such as that adopted under multilateral environmental agreements*? |

**Section D: water, sanitation and health care waste interventions**

**Section A: Demographic data section**

**1.2 Key informant information:**

1.2.1. Age

1.2.2. Gender Male Female

1.2.3 Job description

1.2.4. Years employed in current job:

1.2.5. Highest Qualification:____________________________

1.2.6. Any training on facility climate resilience yes no

1.2.7: if yes provide details______________________________________________________

| Section C: Energy Interventions (Climate Resilience) | | | | |
|---|---|---|---|---|
| **Monitoring and assessment (Climate resilience)** | | | | |
| | **Unavailable; not done** | **In progress; incomplete** | **Completed; achieved** | **Observations** |
| Have you assessed energy needs, availability and alternative sources of renewable energy? | | | | |
| Have you assessed points of greatest heat loss in buildings (such as roofs, especially flat ones) and added or upgraded insulation, and draught proofing*? | | | | |
| Have you periodically checked emergency backup generators, even if rarely used? | | | | |
| Is the renewable energy (such as solar) sufficient to power equipment like refrigerators? | | | | |
| Have you assessed all heating, ventilation and air conditioning ductwork pipes, ensuring they are in good condition and supported adequately by the facility building structure | | | | |
| Have you assessed that location of energy backup or renewable energy infrastructure can withstand extreme weather events (such as strong winds, hail, floods)? | | | | |

| |
|---|
| Does the emergency backup covers at least all critical service areas and equipment? |
| **Risk Management (Climate resilience)** |
| Do you have a plan developed for managing intermittent energy supplies or system failure*? |
| Is there an established maintenance plan to fix easily preventable energy problems? |
| Are there mechanisms in place to filter indoor and ambient air pollutants? |
| Are there combined heat and power systems in place to obtain energy efficiency? |
| Can the energy systems can cope with most extreme weather events*? |
| Are voltage stabilizers available to protect equipment from electrical damage that may be caused by voltage frequency fluctuations (when using a generator), or voltage surges (such as due to power transmission problems in the grid)? |
| **Health and safety regulation (climate resilience)** |
| Has there been updates to building insulation and windows to comply with energy codes? |
| Are emergency electricity generators available to provide required electrical power if the municipal grid, or if the internal normal electrical system fails*? |
| Is critical back-up power supplies available for building infrastructure (such as electrical power, heating and cooling)*? |
| Are solar water heaters available for health care facility's hot water needs? |
| Is there backup energy equipment sufficiently elevated in areas prone to floods and anchored in areas prone to strong winds? |
| Is an adequate backup energy source available if the main |

| |
|---|
| source fails during an extreme weather Event? |
| Is there adequate lighting, communications, refrigeration and sterilization equipment are available during climate related disasters or emergencies? |
| **Monitoring and assessment (Environmental sustainability)** |
| Has the health care facility's energy use and practices (such as percentage of grid-electricity, percentage of fuel oil and liquid gas used)* been assessed? |
| Is there improved training and capacity of health workforce on energy access and performance? |
| Has the health care facility been assessed to determine how and where energy use can be reduced, or increased in energy poor areas? |
| Is the use of air conditioning monitored, and use reduced depending on temperature conditions? |
| **Risk Management (Environmental sustainability)** |
| Have you prioritized energy sources and saving measures which are least costly to introduce and/or those which would bring the biggest saving? |
| Have you installed energy-efficient lighting (such as light emitting diode (LED))? |
| Is natural light used wherever possible? |
| Are you opening windows and making use of natural air flow and light? |
| Have you added occupancy sensor switches for lighting in frequently unoccupied spaces? |
| Have you replaced older air conditioners, refrigerators and other appliances and medical equipment with energy efficient models |
| Is the health care facility fossil fuel consumption reduced by use of renewable energy sources, |

| |
|---|
| including solar (photovoltaic) power, wind power, hydro power and biofuels* |
| Is the diesel-powered generator converted to use biofuels when feasible? |
| Have energy efficient ceiling fans been installed? |
| Are leaks in air conditioning devices plugged? |
| Are the freezers and refrigerators defrosted regularly when required? |
| **Health and safety regulation (climate resilience)** |
| Do you have established education and awareness campaigns to reduce energy use with the participation of all staff? |
| Have you developed system of good practices of energy use conservation with incentives? |
| Have you developed a culture of energy saving by turning off office lights, computers and other equipment, and unplugging electronic devices when not in use? |
| Have you established strategies to lower energy use? |
| Are there design features that maximize natural ventilation such as high ceilings, large windows and skylights (without compromising the structural integrity of the building)? |
| Have you developed an energy management plan to measure energy consumption*? |
| Have you optimized the use of on-site renewable energy? |
| Is there renewable energy powering energy efficient Lighting? |
| Have you implemented a sustainable energy-saving programme in each department? |
| Is there an on-site solar photovoltaic system with battery storage either as a primary or backup electricity source istalled? |
| Is proper maintenance and repair for off-grid |

| |
|---|
| solar photovoltaic power systems* provided? |

**Section E: Infrastructure, Technology and Products Interventions**

**Section A: Demographic data section**

**1.2 Key informant information:**

1.2.1. Age

1.2.2. Gender Male Female

1.2.3 Job description

1.2.4. Years employed in current job:

1.2.5. Highest Qualification:___________________________

1.2.6. Any training on facility climate resilience yes no

1.2.7: if yes provide details____________________________________________________

| Section C: Infrastructure, Technology and Products Interventions (Climate Resilience) | | | | |
|---|---|---|---|---|
| **Adaptation of current systems and infrastructures (Climate resilience)** | | | | |
| | **Unavailable; not done** | **In progress; incomplete** | **Completed; achieved** | **Observations** |
| Are there established partnerships between the health care facility, community and local authorities to reduce climate vulnerability in the surrounding communities*? | | | | |
| Have hazards that can put the health care facility's structural and non-structural elements in danger been assessed? | | | | |
| Have you mapped exposure of health care facility to all types of hazards and risk of the events (such as biological, chemical, geological, hydrometeorological, technological, societal)* | | | | |
| Have you mapped the catchment area of the health care facility in terms of the geographical area and population for whom the health care facility would be expected to provide health care for extreme climate event emergencies and disasters*? | | | | |
| Is the building regularly inspected, both internally | | | | |

| |
|---|
| and externally, for signs of deterioration such as broken plaster, cracks or sinking structural elements, and the causes determined? |
| Does the health care facility has sufficient natural ventilation with protection against disease Vectors? |
| Is the siting of new health care facilities follows assessments to avoid high-risk coastal areas, or areas that are prone to damage from hurricanes, windstorms, floods or water surges, including rising sea-levels associated with climate change*? |
| Are the health care facilities built or retrofitted to cope with extreme weather events ensuring their resilience, safety and continuous operation*? |
| Have you evaluated condition and safety of structural and non-structural elements of the health care facility, resulting from previous exposure to natural and other hazards*? |
| Does new infrastructure construction consider a range of climate related risk scenarios, such as drought, flood, prolonged rainfall, storms, strong winds, heat waves and sea-level rise*? |
| Does the construction and retrofitting of health care facilities follow expert's advice incorporating the topography, flood history and local climate*? |
| Have you assessed health care facility structures and trees along the access routes that would impede traffic if they fell during a climate related emergency or a disaster*? |
| Have you verified that health care facility exit and evacuation routes are clearly marked and free of obstacles to enable emergency evacuation*? |
| Is the health care facility building built with fire resistant and non-toxic materials*? |

| |
|---|
| Has the safety of the location of critical services and equipment in case of flood been assessed? |
| Are glass windows are laminated or otherwise protected to prevent threat from shattering during disasters? |
| Are glass walls, doors and windows able to resist basic wind speeds of 200–250 kph*? |
| Do windows have wind and sun protection devices and are they leakproof? |
| Can power-operated doors be opened manually to permit exit in the event of power failure? |
| Are electrical systems safely secured with backup arrangement to satisfy the facility's demand for at least three days, at all times*? |
| Are information and telecommunications systems safely secured with backup arrangement (via cloud, satellite) to satisfy the facility's demand, at all times*? |
| Have heating, ventilation and air conditioning systems been safely secured with backup arrangement to satisfy the facility's demand for at least three days, at all times? |
| Are there reflective white roofs on buildings installed to reduce heat impacts? |
| Are roofing materials completely and securely fastened, welded, riveted or cemented? |
| Does the roof drainage system have adequate capacity and is it properly maintained? |
| Is the roof leak-proof and insulated? |
| Is there improved safety roofing designed to withstand wind velocity of 175–250 kph in high intensity tropical storm prone areas*? |
| Does the water supply system have sufficient reserves, with backup arrangement, to satisfy the facility's demand for at least three days, at all times*? |

| |
|---|
| Are there sufficient resources allocated for mitigating and preventing climate change impacts of extreme weather events? |
| Are the equipment and supplies (furnishings, medical and laboratory equipment and supplies) safely secured in sufficient quantity and quality with backup arrangement to satisfy the facility's demand for at least three days, at all times*? |
| Is funding is available for newly planned improvements*? |
| **Promotion of new systems and technologies: (Climate resilience)** |
| Is there a national and local early warning system developed for early action to respond to extreme weather events*? |
| Does the health care facility obtains alert information from early warning systems for extreme weather events to ensure prompt action*? |
| Are there plans in place for operating and maintaining critical systems in emergencies and disasters? |
| Is the climate hazard vulnerability analysis prepared and regularly updated (including the impacts of extreme weather risks on infrastructure)*? |
| Has the intensity and probability of extreme weather events across the health care facility (present and future) been mapped*? |
| Have you identified and mapped the health care facility's vulnerabilities and risks to climate-related impacts, emergencies and disasters? |
| Have capacities and resources available within the health care facility to cope with any climate related emergency and disaster been identified? |
| Have health workers been trained to respond to new infectious diseases threats emerging from climate related events or environmentally related, including |

| |
|---|
| post-disaster case management and proper infection prevention and control? |
| Have health information systems with climate information to provide information for early health interventions been strengthened*? |
| Have you ensured that a mechanism exists for the prompt maintenance and repair of equipment required for essential services*? |
| Is the building design responsive to assessment of local hazards*? |
| Are there devices and equipment installed for monitoring indoor temperatures, cooling existing buildings and spaces, blocking direct sun, increasing air flow in case of extreme heat? |
| Is there reliable and sustainable primary and backup communication systems (such as satellite phones, mobile devices, landlines, Internet connections, pagers, two-way radios, unlisted numbers) available including access to an updated contact list for emergency operation*? |
| Are there established mechanisms to identify and incorporate new risks to food supply from climate related impacts*? |
| Does the health care facility use proven smart materials and applications, sensors, low power electronics and similar health care appropriate technology (such as telemedicine, remote sensing systems)*? |
| **Sustainability of health care facility operations (climate resilience)** |
| Are climate related hazards (current and potential) classified as high (indicating a high probability of hazards taking place or high-magnitude hazards, or both), medium (a high probability of moderate hazards) and low (a low probability or hazards of low magnitude)*? |

| |
|---|
| Is there sufficient emergency room surge capacity available to manage climate-related emergencies and disasters (such as extreme heat events)? |
| Is there a disaster risk reduction plan to protect essential services is known and understood by all staff? |
| Is the a health care facility's health emergency plan available for preparedness and response with a clear budget line? |
| Are actions implemented to improve work productivity and financial returns that would otherwise be lost from climate-sensitive health impacts? |
| Are medicines available to cover surge demand to ensure that the health care facilities can sustain the provision of essential and specialized services in an emergency or disaster*? |
| Are essential supplies and pharmaceuticals stockpiled in accordance with national guidelines ensuring timely use to avoid loss due to expiration*? |
| Is there access to antibiotics, antiparasitic and antiviral drugs available for use in acute outbreaks of vector- or water-borne diseases made worse by climate change? |
| Have you estimated the consumption of essential supplies and pharmaceuticals (such as amount used per week) using the most likely extreme weather event Scenarios? |
| Is there an updated inventory of all equipment developed and maintained monthly, including a shortage alert and delivery mechanism? |
| Are there emergency standard operating procedures for extreme weather events includes how and where the health care facility would be evacuated, what disaster recovery steps would be taken to restore some level of services, and how to locate family |

| |
|---|
| members and staff who are off duty at the time*? |
| Are climate related disaster plans regularly updated, and workforce regularly trained on how to implement it? |
| Do you anticipate the impact of the most likely disaster events on the supply of water, food and energy*? |
| Is there a centralized emergency transportation system in place for shifting critically ill patients in case of emergencies or disaster*? |
| Are patient medical records are safely stored particularly in flood-prone areas? |
| Are there established protocols for the health care facility's food service to respond and recover from an extreme weather event (such as emergency menus) and foodborne outbreaks (sanitation, disinfection, isolation)*? |
| Is there secure access to essential backup food sources via multiple agreements with different vendors and through cooperative agreements with other health care facilities*? |
| Are food resources monitored during emergencies to ensure adequate supplies throughout the duration of the event ensuring protocols are in place to guide the rationing of limited food supplies*? |
| Do food service staff adopt proper sanitary food handling and storage? |
| Is there an identified space within the health care facility for the storage and stockpiling of additional supplies, taking ease of access, security, temperature, ventilation, light exposure, and humidity level into consideration? |
| Is there safe access to critical backup supplies and resources are available (for medical equipment, laboratory and treatment supplies, personal protective |

| |
|---|
| equipment, technical experts, alternative energy supplies)? |
| Are there established contingency agreements (such as memoranda of understanding, mutual aid agreements) with vendors to ensure the procurement and prompt delivery of equipment, supplies and other resources in times of shortage*? |
| Have verified measures been taken to protect critical supplies such as emergency power, medicines and patients' records, in case of flood? |
| Are there appropriate backup arrangements available for essential lifelines, including water, power and oxygen*? |
| Are the generator's housing or powerhouse protected from extreme weather events and movable if required? |
| Is there an emergency generator with capacity to meet priority health care facility demands available? |
| Have you ensured an uninterrupted cold chain for essential items requiring refrigeration*? |
| Are there vaccine refrigerators with adequate holdover times available to keep vaccines cool during prolonged periods of power outages? |
| Are there adequate supplies for safe water available (such as chlorine, filters or other water treatment technology, rapid water testing kit, water quality monitoring record sheets)? |
| Have you identified alternative water sources to keep health care facility operational at all times (such as deep well, local water utility, mobile water storage tank)*? |
| Are there mechanisms in place to notify health care facility staff, patients and visitors of air pollution advisories and warnings*? |

| Adaptation of current infrastructures (Environmental sustainability) |
|---|
| Have environmental sustainability criteria been included in health care facility construction or renovation plans*? |
| Are new health care facilities designed and constructed based on low carbon approaches? |
| Is information and operational funds available for energy-saving interventions*? |
| Are medical gases and chemicals stored securely in well ventilated areas? |
| Is the health care facility equipped with air pollution filters to improve indoor air quality? |
| Does construction or retrofitting consider corridors with exterior walls to maximize use of daylight and natural ventilation*? |
| Has retrofitting of buildings been implemented to cut energy waste*? |
| Have solar water heaters been installed? |
| Have hybrid systems (which include renewable energy, batteries, and backup generators) been installed? |
| **Promotion of new technologies (Environmental sustainability)** |
| Has e an energy system according to factors relevant to the facility (such as facility size, level of care, budget, operational cost, resource availability, and geographic location) been selected*? |
| Have you assessed and examined medical equipment to ensure they are energy efficient? |
| Are there appropriate technological devices in place according to energy availability and power (such as chest radiography and magnetic resonance imaging machines need considerable amount power to run)*? |
| Have you replaced oversized air conditioning and ventilation systems for smaller energy efficient models, when feasible? |

| |
|---|
| Are there established partnerships with local government for the installation of off-grid energy systems supply*? |
| Have you performed an inventory of medical and other equipment to understand and determine an estimate of the facility's energy needs? |
| Have you evaluated renewable energy technologies available to power the facility? |
| Are there improved off-grid solar photovoltaic power systems? |
| Have you installed clean and renewable energy sources (such as solar panels, wind turbines and biofuels) for lighting, heat generation, pumping and water heating? |
| Have you installed solar lighting in health care facility car parks? |
| Have you replaced medical devices with more water efficient or energy-efficient models*? |
| Have you replaced dishwashers and laundry machines with those having water-saving functions, whenever possible or when replacements are needed *? |
| Have you substituted mercury-containing thermometers and blood pressure measuring devices for affordable, validated and non-mercury containing device alternatives*? |
| **Sustainability of health care facility operation (Environmental sustainability)** |
| Have you implemented a clear environmentally sustainable procurement policy statement or protocol for all types of products, equipment and medical devices used in the health care facility*? |
| Are health care facility staff trained on effective and efficient procurement practices? |
| Are equipment and supplies purchased from local sources as much as possible, when available*? |

| |
|---|
| Are equipment and supplies purchased giving priority to environmentally friendly products (such as minimal packaging, reusable and recyclable products, avoiding hazardous chemicals and non-degradable plastics)*? |
| Does the health care facility purchase energy-efficient products (medical devices, vehicles, computers)*? |
| Does the health care facility promotes local and sustainable food production*? |
| Have changes been made in health care facility service menus and practices, including limiting the amount of meat and dairy products in meals when appropriate? |
| Is there an on-site garden set up as a means to introduce fresh food in the food service operations? |
| Does the health care facility composts food waste when possible? |
| Does the health care facility plant indigenous trees and plants to obtain health co-benefits, such as the provision of natural shade for patients, staff and visitors during extreme heat events? |
| Does the health care facility surroundings have drought-resistant plants in drought prone areas? |
| Is the drainage for wastewater from health care facility constructed and managed to avoid contamination of the health care setting or the surrounding environment*? |
| Are floor-care products free of zinc, heavy metals, phthalates, glycol ethers and ammonia? |
