## Supplementary File 5 for "Warning system for Extreme weather events, Awareness Technology for Healthcare, Equitable delivery, and Resilience (WEATHER) Project: A mixed methods research study protocol"

| | Year 1 | | | | | | | | | | | | Year 2 | | | | Year 3 | | | | Yr 4 | | | |
|---|---|---|---|---|---|---|---|---|---|---|---|---|---|---|---|---|---|---|---|---|---|---|---|---|
| | M1 | M2 | M3 | M4 | M5 | M6 | M7 | M8 | M9 | M10 | M11 | M12 | Q5 | Q6 | Q7 | Q8 | Q9 | Q10 | Q11 | Q12 | Q13 | Q14 | Q15 | Q16 |
| **Project management and communication** | | | | | | | | | | | | | | | | | | | | | | | | |
| Meetings in Durban, University of Kwazulu-Natal | | | | | | | | | | | | | | | | | | | | | | | | |
| Monthly updates | | | | | | | | | | | | | | | | | | | | | | | | |
| Quarterly meetings | | | | | | | | | | | | | | | | | | | | | | | | |
| **WP1- Evidence gathering** | | | | | | | | | | | | | | | | | | | | | | | | |
| Task 1.1 Systematic review of literature: trends in EWE and infectious disease outbraks | | | | | | | | | | | | | | | | | | | | | | | | |
| Task 1.2 Review of historical environmental health government reports regarding water bodies | | | | | | | | | | | | | | | | | | | | | | | | |
| Task 1.3 Design a pathogen and pollutant management tool | | | | | | | | | | | | | | | | | | | | | | | | |
| D1.1 Systematic review on EWEs and infectious disease in S.A | | | | | | | | | | | | | | | | | | | | | | | | |
| D1.2 **:** Comprehensive framework for monitoring health conditions and types of infections | | | | | | | | | | | | | | | | | | | | | | | | |
| D1.3 Pathogen and pollutant Management tool | | | | | | | | | | | | | | | | | | | | | | | | |
| **WP2- Development and implementation of PWSi** | | | | | | | | | | | | | | | | | | | | | | | | |
| Task 2.1 Open integrated PWSi platform | | | | | | | | | | | | | | | | | | | | | | | | |
| Task 2.2 Design and develop predictive algorithm | | | | | | | | | | | | | | | | | | | | | | | | |
| Task 2.3 Design and launch a dashboard for command and control centre for the PWSi | | | | | | | | | | | | | | | | | | | | | | | | |
| Task 2.4 Test, verify and evaluate the PWSi | | | | | | | | | | | | | | | | | | | | | | | | |
| D2.1 Report of weather and environmental data analysis, gaps and location plans for new sensor clusters | | | | | | | | | | | | | | | | | | | | | | | | |
| D2.2 Health and disease/symptoms outbreak data analysis and data gaps report. | | | | | | | | | | | | | | | | | | | | | | | | |
| D2.3 Rainfall and flood (time, intensity, and duration) AI-predictions algorithm | | | | | | | | | | | | | | | | | | | | | | | | |
| D2.4 Disease outbreak AI prediction algorithm | | | | | | | | | | | | | | | | | | | | | | | | |
| D2.5 A mobile app and a command-and-control dashboard for alert and warning (PWSi) | | | | | | | | | | | | | | | | | | | | | | | | |
| D2.6 PWSi launch and community sensitisation | | | | | | | | | | | | | | | | | | | | | | | | |
| D2.7 PhD thesis in implementing PWSI in SA | | | | | | | | | | | | | | | | | | | | | | | | |
| D2.8 Final Report | | | | | | | | | | | | | | | | | | | | | | | | |
| **WP3- Healthcare systems** | | | | | | | | | | | | | | | | | | | | | | | | |
| **Disease burden, health needs, community experiences** | | | | | | | | | | | | | | | | | | | | | | | | |
| Task 3.1 Development of protocol | | | | | | | | | | | | | | | | | | | | | | | | |
| Task 3.2 Development of research tools | | | | | | | | | | | | | | | | | | | | | | | | |
| Task 3.3 Submission for ethical approval | | | | | | | | | | | | | | | | | | | | | | | | |
| Task 3.4 Obtaining relevant permission | | | | | | | | | | | | | | | | | | | | | | | | |
| Task 3.5 Data collection (secondary) | | | | | | | | | | | | | | | | | | | | | | | | |
| Task 3.6 Data collection Primary | | | | | | | | | | | | | | | | | | | | | | | | |
| Task 3.7 Data management and Analysis | | | | | | | | | | | | | | | | | | | | | | | | |
| D3.1 : First draft report submitted | | | | | | | | | | | | | | | | | | | | | | | | |
| **Assessing Resilience of the Health System** | | | | | | | | | | | | | | | | | | | | | | | | |
| Task 3.8 Development of protocol | | | | | | | | | | | | | | | | | | | | | | | | |
| Task 3.9 Development of research tools | | | | | | | | | | | | | | | | | | | | | | | | |
| Task 3.10 Submission of ethical approval | | | | | | | | | | | | | | | | | | | | | | | | |
| Task 3.11 Obtaining relevant permission | | | | | | | | | | | | | | | | | | | | | | | | |
| Task 3.12 Data collection primary | | | | | | | | | | | | | | | | | | | | | | | | |
| Task 3.13 Data management and Analysis | | | | | | | | | | | | | | | | | | | | | | | | |
| Deliverable: First draft report submitted | | | | | | | | | | | | | | | | | | | | | | | | |
| **Train and mentor healthcare workers on resilient health system and response to EWE** | | | | | | | | | | | | | | | | | | | | | | | | |
| Task 3.14 Development of training material | | | | | | | | | | | | | | | | | | | | | | | | |
| Task 3.15 Compilation of training manuals | | | | | | | | | | | | | | | | | | | | | | | | |
| Task 3.16 Submission of training schedule | | | | | | | | | | | | | | | | | | | | | | | | |
| Task 3.17 Implementation of trainign schedule | | | | | | | | | | | | | | | | | | | | | | | | |
| Task 3.18 Evaluation of training | | | | | | | | | | | | | | | | | | | | | | | | |
| Task 3.19 Mentorship site visits | | | | | | | | | | | | | | | | | | | | | | | | |
| Task 3.20 Mentorship evaluation | | | | | | | | | | | | | | | | | | | | | | | | |
| Deliverable: Workpackage close out report | | | | | | | | | | | | | | | | | | | | | | | | |
| **WP4- Realist Evaluation and Health economics** | | | | | | | | | | | | | | | | | | | | | | | | |
| Task 4.1 One focus group (6-8 people) will be conducted in each of the four communities (Umgababa, Merebank, Isipingo and Port Shepstone) and 5-10 in-depth interviews with key informants (CHW's, health managers) will be held | | | | | | | | | | | | | | | | | | | | | | | | |
| Task 4.2 Outcome assessment baseline and post intervention | | | | | | | | | | | | | | | | | | | | | | | | |
| Task 4.3 Mid point focus groups | | | | | | | | | | | | | | | | | | | | | | | | |

| |
|---|
| Task 4.4 Process evaluation |
| Task 4.5 Overarching SROI |
| D4.1 Outcome assessment |
| D4.2 Feedback to WP2 to optimise PWSI |
| D4.3 Improved preparedness of KZN communities |
| D4.4 Develop theory of change |
| D4.5 Establish the social value ration for PWSI |
| D4.6 PhD thesis in health economics |
