## Supplementary File 6 for "Warning system for Extreme weather events, Awareness Technology for Healthcare, Equitable delivery, and Resilience (WEATHER) Project: A mixed methods research study protocol"

**POST WEATHER project SROI QUESTIONNAIRE FOR PARTICIPANTS**

**PLEASE NOTE:** Your contact details and answers will be treated as strictly confidential, and all data will be anonymised. Only the UKZN, UWS, RCSI and UoP research teams will have access to the information you provide.

**SECTION 1**

**Q1. Name** ________________________________________________

**Q2. Age** ____________

**Q3. Gender**

☐ Male ☐ Female ☐ Prefer not to say

**Q4. Ethnic origin**

☐ Indian/Asian ☐ Black African ☐ Coloured ☐ Mixed ☐ White

☐ Other (please specify) _______________________________

**Q5. Community**: ____________________________________________

**Q6. Occupation**: ___________________________________________

**Q7. Email**: ________________________________________________

**Q8.**

Please rank the following experiences in order of relevance you to, from 1 to 8 (1 = most relevant to you, 8 = least relevant to you) as a result of participating in Weather project.

☐ Explore/ return to crafting

☐ Improve mental health & wellbeing

☐ Improve physical health & wellbeing

☐ Learn something new

☐ Meet new people

☐ Reduce fatigue

☐ Reduce stress and/ or anxiety

☐ Spend time in my community

**Q9. In addition to the above, is there anything else not listed above that you did experience from participating in the WEATHER project?**

**The following questions ask about the value you place on having the Early Warning System (EWS) in your local community**

**Consider the following 'hypothetical' situation:**

Suppose that the WEATHER project EWS was no longer available but there was new predictive EWS project available in your local area and was available free of charge.

1. Would you be willing to participate in this alternative EWS project?

☐Yes ☐No

2. Now suppose this new alternative EWS project could no longer be offered free of charge and suppose it was not available through the Department of health/SA Weather service or partner organisations. What is the **maximum amount** that would you be willing to pay at your own expense monthly? **Please consider what you could realistically afford to pay given your current financial situation.**

**Payment ladder in South African ZAR**

| | | | | | | | | | | | | |
|---|---|---|---|---|---|---|---|---|---|---|---|---|
| ☐ R 2.38 | ☐ R24 | ☐ R71 | ☐ R119 | ☐ R238 | ☐ R357 | ☐ R477 | ☐ R596 | ☐ R715 | ☐ R834 | ☐ R952 | ☐ R1072 | ☐ R1191 |
| ☐ Nothing | | | | | | | | | | | | |

3. **These questions have asked about your 'willingness to pay' for hypothetical programs. Below are some sentences to explain why you gave the answers you have chosen. Please read each and select the one(s) that best explain your answers:**

☐ They are the value I would put on the predictive EWS Weather project.

☐ I am not interested in this alternative predictive EWS project described.

☐ I cannot afford any additional tax.

☐ I do not believe the alternative EWS predictive project described.

☐ The government should provide the benefits described by EWS predictive project the without any additional cost for taxpayers

4. **Thinking about the cost of living as it affects you and your household, which of these best, describes your situation at present?**

☐ I find it a strain to get from week to week ☐ I have to be careful about money

☐ I am able to manage without much difficulty ☐ I am quite comfortably off

**Thank you for completing this questionnaire.**

**Your participation in this study is much appreciated.**

Please note that your contact details and answers will be treated as strictly confidential, and all data will be anonymised. Only the UKZN, UWS, RCSI and UoP research teams will have access to the information you provide.
